# A living systematic review, meta-analysis, and open data resource of trials of MDMA-assisted therapy for PTSD

**DOI:** 10.64898/2026.03.27.26349536

**Authors:** Brooke L. Sevchik, S. Parker Singleton, Analiese Lahey, Pim Cuijpers, Mathias Harrer, Megan T. Jones, Sandeep M. Nayak, Eric C. Strain, Simon N. Vandekar, David B. Yaden, Robert H. Dworkin, J. Cobb Scott, Theodore D. Satterthwaite

**Affiliations:** Penn Lifespan Informatics and Neuroimaging Center, University of Pennsylvania, PA, USA; Department of Psychiatry, Perelman School of Medicine, University of Pennsylvania, PA, USA; Department of Clinical, Neuro and Developmental Psychology, WHO Collaborating Centre for Research and Dissemination of Psychological Interventions, Vrije Universiteit Amsterdam, The Netherlands; Babeș-Bolyai University, Ciuj-Napoca, Romania; Amsterdam Public Health Research Institute, Amsterdam University Medical Centre, Amsterdam, The Netherlands; Department of Biostatistics, Vanderbilt University Medical Center, Nashville, TN, USA; Center for Psychedelic and Consciousness Research, Department of Psychiatry and Behavioral Sciences, Johns Hopkins University School of Medicine, Baltimore, Maryland, USA; Department of Psychiatry and Behavioral Sciences, Johns Hopkins University School of Medicine, Baltimore, Maryland, USA; Departments of Anesthesiology and Perioperative Medicine and Neurology, University of Rochester School of Medicine and Dentistry, New York, Rochester, USA; Department of Psychiatry, Brain Behavior Laboratory, Perelman School of Medicine, University of Pennsylvania, Philadelphia, PA, USA; VISN4 Mental Illness Research, Education, and Clinical Center at the Crescenz VA Medical Center, Philadelphia, PA, USA; Penn-CHOP Lifespan Brain Institute, Perelman School of Medicine, Children’s Hospital of Philadelphia Research Institute, Philadelphia, PA, United States

**Author notes:** These authors contributed equally as first authors (Brooke L. Sevchik, S. Parker Singleton). These authors contributed equally as senior authors (J. Cobb Scott, Theodore D. Satterthwaite). **Corresponding author:** Email address (Theodore D. Satterthwaite). Email address (S. Parker Singleton), Address: 3700 Hamilton Walk, 5A, Philadelphia, PA 19104.

## Abstract

3,4-methylenedioxymethamphetamine (MDMA) has emerged as a potential treatment for post-traumatic stress disorder (PTSD), generating considerable enthusiasm in the field. However, rapidly changing evidence in a fast-moving field can be challenging to integrate. Here, we present a living systematic review and open-data meta-analytic resource on MDMA treatment for PTSD. In this initial release, six randomized controlled trials comprising 286 participants are included in the database. Our primary model uses inverse-variance random-effects meta-analysis of standardized mean differences on primary outcomes of PTSD. Compared to control conditions, MDMA showed a greater reduction in PTSD symptoms (Hedges’ *g* = -0.71). Meta-regression on both the number of dosing sessions and cumulative dose showed that a higher number of dosing sessions and a higher cumulative dose was related to larger effects of MDMA. Treatment with MDMA as compared to placebo also resulted in higher response (risk ratio (*RR*) = 1.35) and remission (*RR* = 2.25) rates. Most studies included in the database had a low risk of bias according to Cochrane guidelines, though these fail to capture pertinent challenges in the field such as expectancy, functional unblinding, potential issues with study conduct, and safety. The current findings were assigned an overall low certainty rating using the GRADE approach. Together, this systematic review and meta-analysis suggests that MDMA-assisted therapy results in short-term decreases in PTSD symptoms across studies to date, though more trials are needed. This living systematic review, meta-analysis, database, and online dashboard (sypres.io) will continue to be updated as evidence emerges, providing a valuable, open, and transparent resource for researchers in a rapidly evolving field.

## Introduction

Post-traumatic stress disorder (PTSD) is a major burden for both individuals and society. Patients with PTSD have higher rates of suicide (Demesmaeker et al., 2025; Fox et al., 2021; Wilcox et al., 2009), reduced quality of life (Doctor et al., 2011; Miller et al., 2024), and higher all-cause mortality (Giesinger et al., 2020; Nilaweera et al., 2023). Globally, it is estimated that PTSD is responsible for $23.2 billion a year in health care costs and diminished productivity (Davis et al., 2022; Stanicic et al., 2025). While approximately 6% of the general population experiences PTSD during their lifetime (Koenen et al., 2017), rates are markedly elevated among populations with higher trauma exposure, including military service members (17.1%) and emergency responders (10-32%) (Hoge et al., 2004; Javidi & Yadollahie, 2012).

PTSD is characterized by alterations in arousal and reactivity, cognition and mood deficits, (VanElzakker et al., 2018), re-experiencing symptoms (Ehlers, 2010), and avoidance of trauma-related cues. At present, only paroxetine and sertraline, both selective serotonin-reuptake inhibitors (SSRIs), are approved by the United States Food and Drug Administration (US FDA) to treat PTSD. However, the clinical effectiveness of SSRIs is modest (Akiki & Abdallah, 2018). Psychological treatments are typically considered first-line treatments for PTSD (Hamblen et al., 2019). Trauma-focused approaches such as prolonged exposure therapy and cognitive processing therapy show consistent reductions in PTSD symptoms (Cuijpers et al., 2024). However, these treatments are prone to high drop-out rates (i.e., 20-30%) (Imel et al., 2013; Najavits, 2015; Rutt et al., 2018; Sciarrino et al., 2020), limited efficacy in certain populations (Sripada et al., 2019; Varker et al., 2021), and lack of remission despite significant symptom improvement (Steenkamp et al., 2015). To address this gap, recent studies have examined pharmacological agents that may enhance psychotherapeutic processes (Feduccia et al., 2018), including 3,4-methylenedioxymethamphetamine (MDMA). MDMA has been proposed to attenuate fear responses during traumatic memory recall, potentially reducing avoidance and reactivity symptoms, thereby facilitating patients’ ability to engage with traumatic material (Mithoefer et al., 2011).

The Multidisciplinary Association for Psychedelic Studies (MAPS), and more recently their for-profit spin-off Resilient Pharmaceuticals (formerly Lykos Therapeutics), has developed and evaluated an MDMA-assisted therapy (MDMA-AT) program for PTSD. The typical MDMA-AT protocol involves a co-therapist team delivering 2-3 preparatory psychotherapy sessions spaced approximately one week apart before MDMA dosing, and 2-3 dosing sessions lasting 8-10 hours spaced approximately one month apart, along with 2-3 integration psychotherapy sessions spaced approximately one week apart following each dosing session (Lewis & Byrne, 2023; Reardon, 2024). Their data have shown promise for the efficacy of MDMA: two phase 3 studies, for example, showed a significant mean decrease in the CAPS-V score compared to the placebo group (Mitchell et al., 2021, 2023).

However, this promise must be contextualized within the limitations of current trials. For one, aspects of the psychotherapy design are broad and vague. Study manuals mention potential spontaneous use of both evidence-based techniques and those lacking empirical evidence (Yehuda et al., 2026). The degree to which the non-directive therapy design incorporates each of these elements is largely up to each individual therapist team’s interpretation of the study manual for each participant. This creates the possibility that the psychotherapy model in the placebo group might, in practice, differ significantly from the psychotherapy model used in the intervention group (Cristea et al., 2024). Furthermore, this problem is exacerbated by the high potential for functional unblinding, in which therapist teams often correctly guess each participant’s group assignment due to psychoactive effects of MDMA (van Elk & Fried, 2023). Lastly, the careful selection of participants and small sample sizes limit external validity in these studies.

Given these issues surrounding existing treatments and the challenges in trial design, the need to synthesize evidence for the efficacy of MDMA in treating PTSD symptoms is clear. In this context, the number of systematic reviews and meta-analyses on this topic has grown rapidly (Bahji et al., 2020, 2025; Brett & Bynum, 2025; Højlund et al., 2025; Hood et al., 2024; Hoskins et al., 2021; Illingworth et al., 2021; Kisely et al., 2023; Shahrour et al., 2024; Smith et al., 2022; Stanicic et al., 2025; Sze Jing Yong et al., 2025; Yang et al., 2024; Žuljević et al., n.d., 2025). However, in a fast-moving field these meta-analyses quickly become outdated, failing to capture the most recent findings (Iversen & Quintana, 2026). Looking forward, there are already thirteen active phase 2 or phase 3 trials evaluating MDMA for PTSD (Search ClinicalTrials.Gov For, n.d.), data from which are not included in existing meta-analyses. In addition, prior meta-analyses lack transparency and reproducibility, often lacking raw data, code, and clear justifications for analytic decisions.

Living systematic reviews (LSRs) can help address these challenges and considerations, as they incorporate continuous updates as the evidence base grows and shifts (Simmonds et al., 2022). LSRs are especially useful when the certainty of the evidence base is low (Iversen & Quintana, 2026). The “living” nature of these reviews automatically imbues added caution when interpreting results from existing data, as the dynamic nature of the evidence base introduces the possibility that results could change significantly as data from more studies are added (Simmonds et al., 2022).

Accordingly, here we present a living systematic review and meta-analysis on MDMA treatment for PTSD symptoms, ensuring that the latest findings are reflected in the data synthesis. In addition, our continuously maintained database of raw data and effect sizes is released using the Metapsy infrastructure (metapsy.org), and are summarized in an interactive online dashboard (metapsy.org/sypres/mdma-ptsd). This dashboard allows scientists, clinicians, and the broader public to retrieve the current evidence on the efficacy of MDMA for PTSD. Moreover, it allows individuals to examine the influence of inclusion criteria, analysis choices, and individually-selected subgroups on results. Lastly, all of our analysis code and data is publicly available, adhering to modern standards of reproducible science. Herein, we describe our initial synthesis of these data. We plan to update our review at least annually; all data, code, and results are available on our public website: SYPRES (Synthesis of Psychedelic Research Studies; sypres.io).

## Methods

This meta-analysis is a pre-registered study under the SYPRES project, and the study protocol is available on PROSPERO (CRD42024584945). Please refer to the Supplementary Information for detailed methodological information.

### Eligibility Criteria

We included randomized controlled clinical trials published in English in peer-reviewed journals comparing MDMA for PTSD with a comparator in adult (>18 years old) populations. In order to meet these criteria, the study population needed to include individuals with elevated PTSD symptoms; studies with only healthy participants were not considered. Eligible interventions included any dose and formulation of MDMA or other prodrugs of MDMA intended to produce an alteration of subjective experience in the patient, with or without the conjunctive use of therapy (e.g., microdosing studies were not included). Eligible comparators included any form of placebo with or without the conjunctive use of therapy, including lower doses of the intervention drug and any dose of other psychotropics intended to improve blinding (without known therapeutic efficacy for PTSD). Eligible comparators also included control for spontaneous improvement via waitlist or usual care. Additionally, for studies that used a crossover design, data from pre-crossover timepoints was required to account for the potential carry over effects of an MDMA dose.

### Study search and selection

We searched PubMed, Embase, PsycInfo, Web of Science, Scopus, and the reference lists of systematic reviews retrieved from the searches. Expert research librarians assisted in crafting study search criteria. The final search was conducted on November 25, 2025 (see the Supplementary Information for search terms). Study screening and data extraction was independently performed by two reviewers. Two reviewers assessed risk of bias (RoB) using Cochrane’s Risk of Bias 2.0 tool (see the Supplementary Information for details). We rated the certainty of the evidence synthesized in this review using the GRADE approach (Guyatt et al., 2008). We plan to update our search and the accompanying analyses and online resources at least annually for the next five years. This study’s senior authors will be responsible for on-going supervision.

### Effect size calculation

We used each study’s endpoint sample sizes, means, and standard deviations for treatment and control to calculate Hedges’ *g* for each study as the primary effect size measure for continuous outcomes. Hedges*’ g* is a small-sample bias corrected standardized mean difference (SMD). SMD is frequently used as a summary statistic in meta-analysis when different measurements are used across studies to assess the same outcome (e.g., different versions of the Clinician Administered PTSD scale in this case). SMD standardizes results from different measures to a uniform scale by expressing the size of the intervention effect relative to between-participant variability under the assumption that differences in standard deviations across studies are only due to differences in the measurement scales and not real differences in variability among the populations studied (Higgins et al., 2024). For dichotomous outcomes, we calculated risk ratios (RRs) from raw event data. RRs quantify how much an intervention multiplies the chance of an outcome occurring. For instance, when an intervention has an RR of 3, this indicates that the outcome is three times more probable in the intervention arm than the control arm. Likewise, an RR of 0.25 means that the probability of an outcome occurring in the intervention arm is one-fourth that in the control arm. A RR of 1 indicates equal probabilities in both arms. Therefore, RRs provide an easily interpretable summary statistic of dichotomous outcomes (Higgins et al., 2024). See *Selection of effect sizes* in the Supplementary Information for more details.

### Meta-analyses

#### Primary analysis on continuous outcomes

We performed inverse-variance random-effects modeling of SMDs on primary outcomes of PTSD symptoms at the primary endpoint specified in the original study publication. For three-arm trials, we compared the high-dose arm to the low-dose control arm to maximize the contrast in dose exposure between the intervention and control groups. While the outcomes were similar within the small number of studies included in the meta-analysis, a random-effects model was chosen to account for potential sources of between-study variability in the true effect. Potential sources of this variability arise from differences in dose amount and dosing schedules, different control group comparators, different versions of the PTSD scales used to assess symptoms, and different primary endpoints. See the Supplementary Information for tables and procedures detailing the selection of primary outcomes and timepoints for studies that reported multiple outcome measures and/or multiple timepoints during which an outcome was assessed. Between-study heterogeneity (*tau**^2^***) was calculated using the Restricted Maximum Likelihood (REML) estimator (Viechtbauer, 2005) and the Q-profile method to calculate the confidence interval (Viechtbauer, 2007). We employed the Knapp-Hartung adjustment to calculate the confidence interval for the pooled effect size (Knapp & Hartung, 2003). This adjustment creates a more conservative confidence interval and *p*-value that varies less with changes in heterogeneity variance. Funnel plots and Egger’s test were used to assess small study bias, although the small number of studies limits the precision of these approaches (Egger et al., 1997).

#### Three-level correlated and hierarchical effects meta-analysis

To assess MDMA’s effects independent of measurement timepoint or dose, we applied a three-level meta-analysis model on 15 effect sizes from the six studies included in our database. Effect sizes from these studies were calculated at timepoints ranging from 4 to 18 weeks after baseline, and from 1 to 3 dosing sessions. These effect sizes included comparisons of both high and medium-dose arms against low-dose arms for three-arm trials. These effect sizes were limited to assessments on the primary instrument occurring at least 1 day after dosing and before any crossover between groups. See **SI Table 4** in the Supplementary Information for full description of each effect size. Variance-covariance matrices of each study with two or more effect sizes were estimated using a constant within-study correlation coefficient (*ρ*) of 0.6, creating a “correlated and hierarchical effects” (CHE) model. Cluster-robust variance estimation was used to guard against potential model misspecification. Heterogeneity was calculated using the REML estimator with parametric bootstrapping (5,000 iterations) used to generate confidence intervals around the heterogeneity variance components. To examine how the number of dosing sessions and the cumulative dose influence effect size, we additionally performed two separate linear meta-regressions using the three-level CHE model: (a) adding the number of dosing sessions as a continuous predictor in one case, and (b) adding the cumulative dose in the intervention arm as a continuous predictor in the other. Cumulative dose was defined as the cumulative dosage of MDMA administered across the sessions prior to the outcome assessment.

#### Meta-analysis on dichotomous outcomes

In addition to continuous outcomes, we evaluated dichotomous response (*k* = 5) and remission (*k* = 4) outcomes reported by studies in our database. Response typically refers to the minimum change in a scale to signify clinically relevant improvement in symptoms, while remission typically refers to an endpoint measurement that falls below a specified cutoff for a diagnosis of PTSD. Both of these variables were defined and reported by each individual study. Response definitions range from a reduction in CAPS severity from severe to mild, a decrease of more than 30% in CAPS total score, or a decrease of at least 10 points on the CAPS. Remission definitions encapsulate a loss of PTSD diagnostic criteria on the CAPS, with some also specifying the need for a CAPS total severity score of 11 or less. We used inverse-variance random-effects modeling of log-transformed risk ratios, which were retransformed after pooling. Between-study heterogeneity was calculated with the Paule-Mandel (PM) estimator (Paule & Mandel, 1982), which is an alternative to REML with a good performance in analyzing dichotomous outcomes. Confidence intervals were again calculated using the Q-profile method for heterogeneity estimates and the Knapp-Hartung adjustment for pooled effect sizes.

#### Meta-analysis on co-morbid depression symptoms in PTSD

We also assessed MDMA’s effects on co-morbid depression symptoms in the three studies that reported outcomes on depression instruments. In each study, the BDI-II instrument was the only depression instrument reported. We calculated SMDs using BDI-II at the primary endpoint and performed inverse-variance random-effects modeling using the same methodological parameters as our primary analysis on continuous outcomes.

#### Sensitivity analyses

To assess the robustness of our primary findings, the following sensitivity analyses were also performed:

1. Fixed effect models: We ran fixed/common-effect models as sensitivity analyses to compare to random effects models. Fixed effect models assume that the between-study variance (*tau**^2^***) is 0, such that all studies share a common true effect size. For our continuous model, we used a standard inverse-variance weighting fixed-effect model on standardized mean differences (Hedges’ *g*). For our dichotomous models, we used a standard inverse-variance weighting fixed-effect model on the log risk ratio.
2. Alternate dosing in three-arm trials: Given that Mithoefer 2018 and Ot’alora G 2018 employed a three-arm design comparing MDMA at high and medium doses against a low-dose control, we conducted a sensitivity analysis substituting the medium dose intervention arm for the high-dose intervention arm used in our primary analysis.
3. Three-level model within-study correlation coefficient sweep: Our three-level CHE model assumed a constant within-study correlation coefficient (ρ) of 0.6, which is typically a good approximation for datasets with unknown and/or complex dependence structures (Pustejovsky & Tipton, 2022). To test the sensitivity of our results against this approximation, we recalculated Hedges*’ g* as a function of *ρ* from 0 to 1 in 0.1 increments.
4. Bayesian meta-analysis: We replicated our primary meta-analysis on continuous outcomes using a Bayesian implementation. We used “weakly informative” prior distributions for both the main effect and the heterogeneity parameter *tau* that have been recommended by prior work (Röver et al., 2021; Williams et al., 2018). The main effect prior was a normal distribution centered around 0, with a standard deviation of 1, while the *tau* prior was a half-normal distribution with a standard deviation of 0.5.

### Software, data, and code availability

Literature screening and data extraction was performed using Covidence. Meta-analyses were conducted using R (4.4.1) in RStudio (2024.04.2+764) with metapsyTools (1.0.13) (Harrer et al., 2022), a package of helper functions for Metapsy that uses meta (7.0.0)(Balduzzi et al., 2019), metafor (4.6.0) (Viechtbauer, 2010), and dmetar (0.1.0) (Harrer et al., 2019) functions. The Bayesian meta-analysis was implemented using the bayesmeta (3.5) (Röver, 2020) R package. The living database used for this analysis can be accessed through Metapsy (docs.metapsy.org/databases/ptsd-mdmactr/) or downloaded from our website, where code for all analyses is also openly available (sypres.io).

## Results

We identified 1,586 reports from our searches. Of these, 26 passed initial title and abstract screening and were reviewed as full-text articles. Six studies (Mitchell et al., 2021, 2023; Mithoefer et al., 2011, 2018; Oehen et al., 2013; Ot’alora G et al., 2018) met inclusion criteria for our primary model. Complete information about study identification and screening is included in **Fig. 1**. Our database consists of 61 effect sizes generated from these 6 included studies, covering continuous and dichotomous measures of PTSD symptoms across timepoints (from four days after the first dose out to 12 months post-baseline). We released this database publicly on Metapsy (P. Singleton et al., 2026); it served as the basis for all following analyses. All studies included a psychotherapy or psychological support component before, during, and after MDMA dosing (see *Psychotherapy or Psychological Support* in the Supplementary Information). See **Table 1** for detailed study characteristics. Overall, of the six studies in the database, one study had some concerns (Mithoefer 2011), and five studies were deemed to have an overall low risk of bias (see **Table 2** for greater details). Mithoefer 2011 had some concerns due to what appears to be a per-protocol study design, where dropouts were replaced with new enrollees. According to Cochrane guidelines, per-protocol study designs are not an appropriate analysis to estimate the effect of assignment to intervention.

**Fig. 1:**
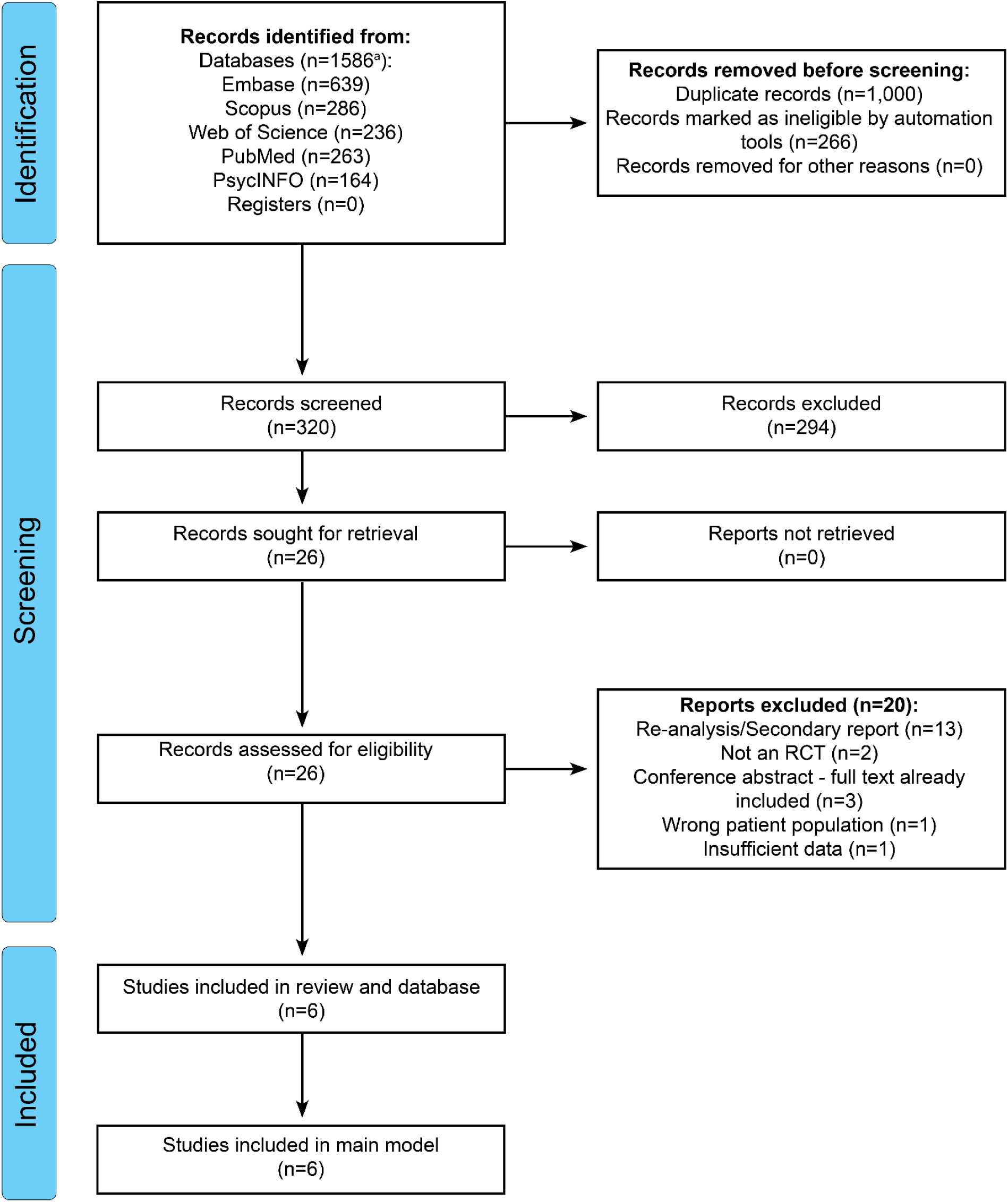
PRISMA 2020 flow diagram. ^a^Includes 2 records that were merged with their parent studies from a total of 1588 original records; see the Supplementary Information for more details

**Table 1:** Summary of RCTs on MDMA for PTSD symptoms. N given is the number of participants randomized. The intervention columns of this table exclude the medium-dose arms for Mithoefer 2018 and Ot’alora 2018 (*n* = 7, 75 mg MDMA; *n* = 9; 100 mg MDMA, respectively). All studies included psychotherapy in both arms. In all studies, participants were eligible to receive a supplemental dose equivalent to half of the initial dose partway through dosing sessions. Study endpoints are the reported primary endpoints for each study and given in weeks since the final dose. Summary statistics across studies (blue rows) were calculated as weighted averages. US = United States; CH = Switzerland; Multi = multi-site. Mitchell 2021 was conducted at sites across the US, Canada, and Israel. Mitchel 2023 was conducted at sites in the US and Israel. PTSD = Post-traumatic stress disorder.

| Study ID | N | Country | Age (SD) | Female (%) | Previous MDMA use (%) | Patient diagnosis | Intervention | Control | Primary Endpoint |
| --- | --- | --- | --- | --- | --- | --- | --- | --- | --- |
| Mithoefer 2011 | 23 | US | 40.4 (7.2) | 85 | 43 | PTSD | Two doses MDMA (125 mg) | Two doses placebo | 8 weeks |
| Oehen 2013 | 14 | CH | 41.4 (11.2) | 83 | 8.33 | PTSD | Three high doses MDMA (125 mg) | Three low doses MDMA (25 mg) | 3 weeks |
| Mithoefer 2018 | 26 | US | 37.2 (10.3) | 27 | NA | PTSD | Two high doses MDMA (125 mg) | Two low doses MDMA (30 mg) | 4 weeks |
| Ot'alora 2018 | 28 | US | 42.0 (12.9) | 67.9 | NA | PTSD | Two high doses MDMA (125 mg) | Two low doses MDMA (40 mg) | 4 weeks |
| Mitchell 2021 | 91 | Multi | 41.0 (11.9) | 65.6 | 32.2 | PTSD | Three doses MDMA (80; 120; 120 mg) | Three doses placebo | 9 weeks |
| Mitchell 2023 | 104 | Multi | 39.08 (10.34) | 71.19 | 46.16 | PTSD | Three doses MDMA (80; 120; 120 mg) | Three doses placebo | 9 weeks |
| **Total N** | **286** | - | - | - | - | - | - | - | - |
| **Mean** | **47.67** | - | **40.03** | **66.76** | **38.59** | - | - | - | **7.68 weeks** |

**Table 2:** Additional study characteristics and risk of bias assessments using Cochrane’s Risk of Bias 2.0 tool. Randomization = bias due to randomization process; Deviations = bias due to deviations from intended interventions; Missingness = bias due to missing outcome data; Measurement = bias due to measurement of the outcome; Selection = bias due to selection of the reported results; Overall = overall risk of bias; CAPS-IV = Clinician-Administered PTSD Scale (v4); CAPS-V = Clinician-Administered PTSD Scale (v5); PDS = Posttraumatic Diagnostic Scale (PDS).

| Study ID | Design | Scale | Assessor | Risk of Bias Domains |  |  |  |  | Overall |
| --- | --- | --- | --- | --- | --- | --- | --- | --- | --- |
|  |  |  |  | Randomization | Deviations | Missingness | Measurement | Selection |  |
| Mithoefer 2011 | Double-blind placebo-controlled | CAPS-IV | Independent study staff | Low | Some concerns | Low | Low | Low | Some concerns |
| Oehen 2013 | Double-blind placebo-controlled | CAPS-IV | Independent study staff | Low | Low | Low | Low | Low | Low |
| Mithoefer 2018 | Double-blind placebo-controlled | CAPS-IV | Independent study staff | Low | Low | Low | Low | Low | Low |
| Ot'alora 2018 | Double-blind placebo-controlled | CAPS-IV | Independent study staff | Low | Low | Low | Low | Low | Low |
| Mitchell 2021 | Double-blind placebo-controlled | CAPS-V | Third-party | Low | Low | Low | Low | Low | Low |
| Mitchell 2023 | Double-blind placebo-controlled | CAPS-V | Third-party | Low | Low | Low | Low | Low | Low |

### MDMA treatment significantly reduces PTSD symptoms compared with control conditions

The analysis on continuous outcomes for the six studies included in the primary model showed a statistically significant reduction in PTSD scores after MDMA treatment compared with control conditions (**Fig. 2**; Hedges’ *g* = -0.71 [-0.95; -0.47], *p* < 0.001, *k* = 6, *n* = 242), with low between-study heterogeneity (*tau^2^* = 0.00 [0.00; 0.38], *I^2^*= 0.0% [0.0%; 74.6%]). Visual inspection of a funnel plot (**SI Fig. 1**) revealed limited asymmetry. An Egger’s test did not find small study effects (*intercept* = -0.52 [-1.94; 0.91], *t* = -0.71, *p* = 0.52), although this test is underpowered given the small number of studies included in our meta-analysis (*k* < 10).

**Fig. 2:**
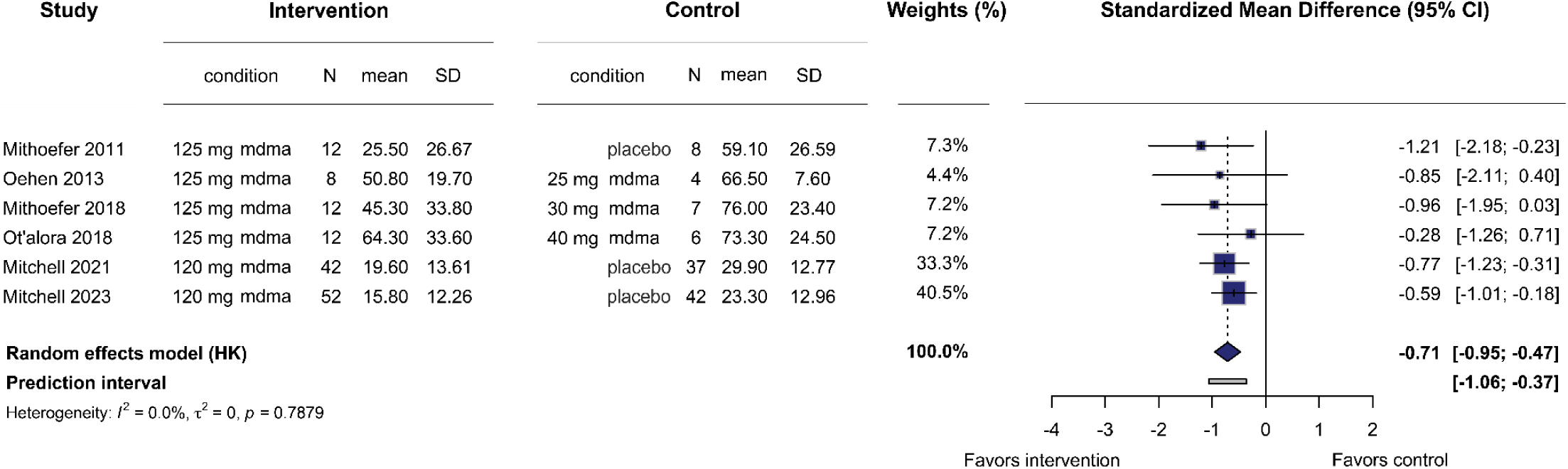
Primary meta-analysis on continuous outcome variables reveals MDMA treatment significantly reduces PTSD symptoms compared with control. Boxes represent the standardized mean difference (Hedges’ *g*) for each study, and the lines extending from the box represent the 95% confidence interval around each effect size, while the size of each box is proportional to its weight. The diamond at the bottom represents the pooled effect size (meta-analytic mean). The gray line at the bottom represents the prediction interval of the expected range of true effects in a new study. HK = Knapp-Hartung adjustment.

### MDMA’s effects increase over dosing sessions and cumulative dose

Our three-level CHE model revealed a significant decrease in PTSD scores with MDMA compared to the control conditions consistent with our primary model (Hedges’ *g* = -0.60 [-0.86; -0.34], *p* < 0.001, *k* = 6, *n* = 286, *tau^2^* = 0.00 [0.00; 0.23], *I^2^* = 0.0% [0.0%; 66.0%]) (**Fig. 3a**). We next performed two separate meta-regressions. First, adding the number of dosing sessions as a continuous predictor to our model (**Fig. 3b**) revealed a significant effect of increased dosing sessions, such that more dosing sessions resulted in a greater between-group difference in PTSD scores (*β* = -0.20 Hedges*’ g* [-0.34; -0.06], *p* = 0.01). Second, we evaluated the cumulative MDMA dose as a continuous predictor; this revealed a significant effect of increased MDMA dose, such that a higher dose exposure resulted in a greater between-group difference in PTSD scores (*β* = -0.002 *Hedges’ g/mg* [-0.003; -0.0005], *p* = 0.01).

**Fig. 3:**
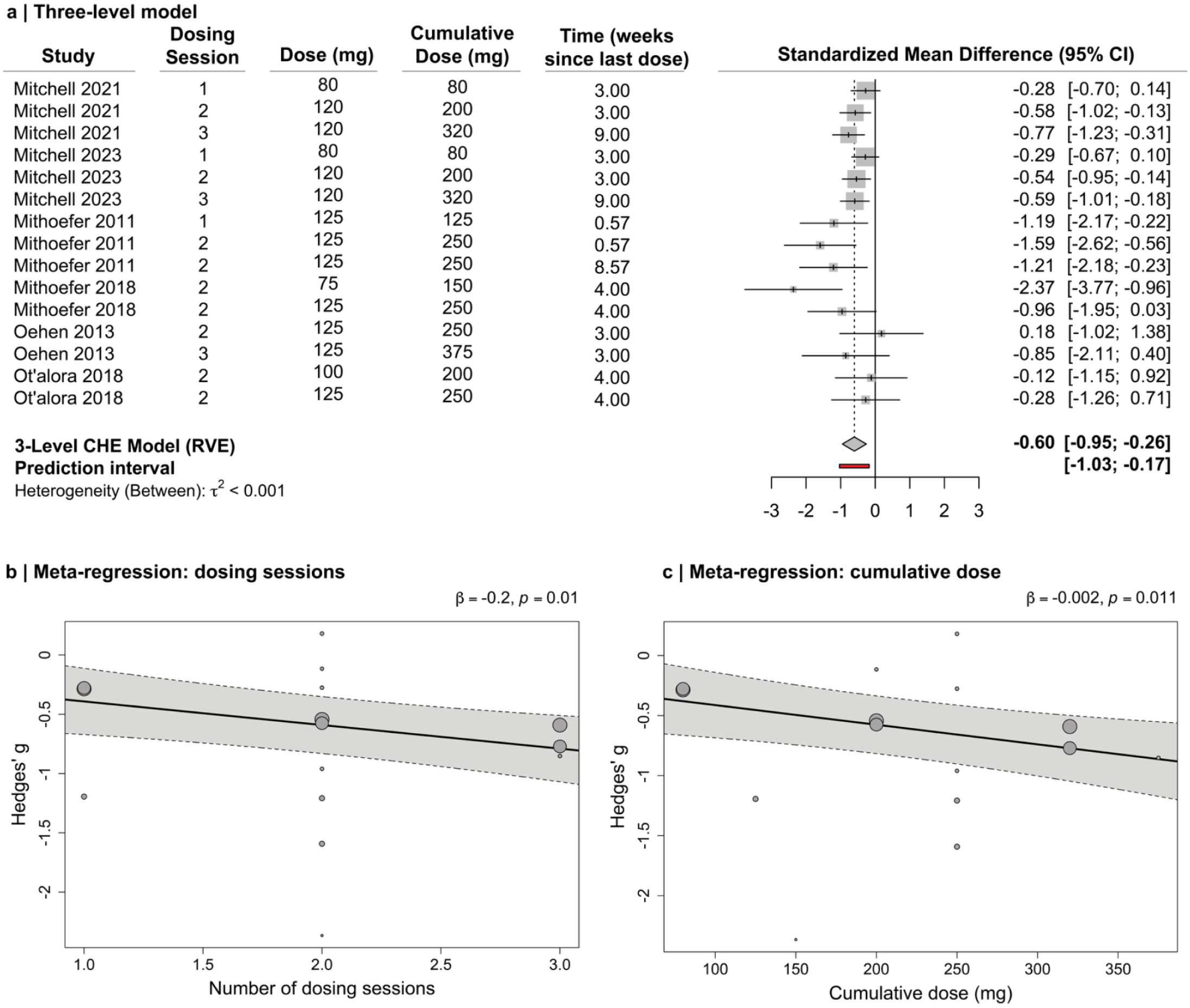
Three-level model and meta-regressions. **a)** Three-level model. Boxes represent the standardized mean difference (Hedges’ *g*) for each study, and the lines extending from the box represent the 95% confidence interval around each effect size, while the size of each box is proportional to its weight. The diamond at the bottom represents the pooled effect size (meta-analytic mean) from the 3-level CHE model with robust variance estimation. The red line at the bottom represents the prediction interval of the expected range of true effects in a new study. RVE = Robust Variance Estimation. **b)** Meta-regression on number of dosing sessions to test dose-response relationship. Individual points represent individual study arms, where the size of each point is proportional to its weight. The solid black line is the fitted meta-regression line, and the shaded region around the solid black line is the 95% confidence interval for the regression. **c)** Meta-regression on cumulative dose to test dose-response relationship. Individual points represent individual study arms, where the size of each point is proportional to its weight. The solid black line is the fitted meta-regression line, and the shaded region around the solid black line is the 95% confidence interval for the regression.

### Secondary outcomes: greater treatment response and remission rate, nonsignificant effect on depression symptoms after MDMA treatment

We next evaluated the impact of MDMA on dichotomous response and remission outcomes. We found evidence for statistically significant greater treatment response with MDMA compared with control conditions (**Fig. 4a**; *RR* = 1.35 [1.10; 1.66], *p* = 0.016, *k* = 5, *n* = 222), with low between-study heterogeneity (*tau^2^* = 0.00 [0.00; 1.68]; *I^2^* = 0.0% [0.0%; 79.2%]). We also found that there were significantly higher remission rates with MDMA compared with control conditions (**Fig. 4b**; *RR* = 2.25 [1.04; 4.87], *p* = 0.044, *k* = 4, *n* = 210), with low between-study heterogeneity (*tau^2^* = 0.00 [0.00; 5.78]; *I^2^* = 0.0% [0.0%; 84.7%]). In contrast, we did not find evidence for a significant effect of MDMA on depression symptoms (**Fig. 4c**; Hedges’ *g* = -0.66 [-2.70; 1.39], *p* = 0.30), with a low number of studies (*k* = 3, *n* = 118) and high heterogeneity (*tau^2^* = 0.44 [0.00; 29.60], *I^2^* = 68.0% [0.0%; 90.7%]).

**Fig. 4:**
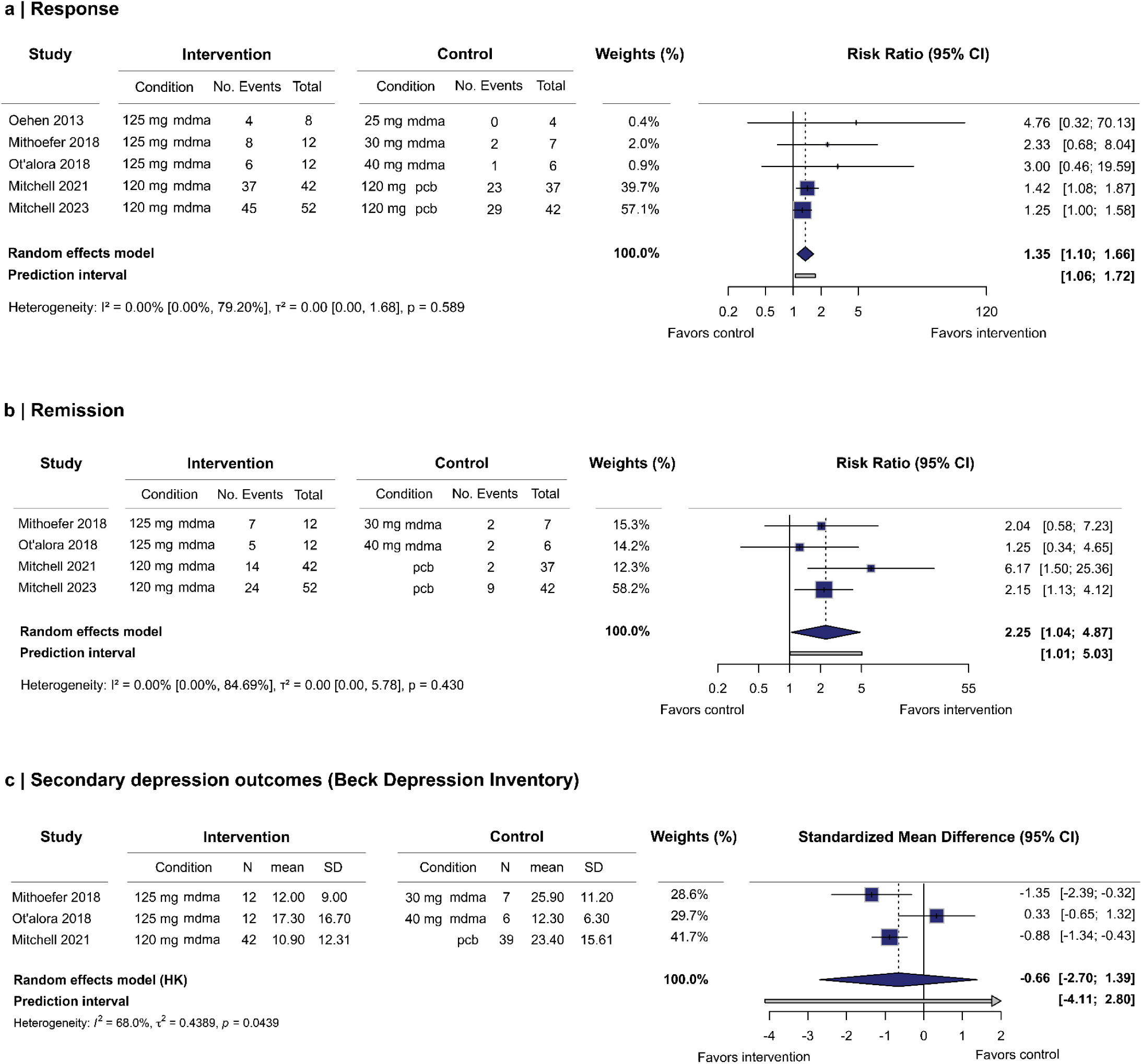
Secondary meta-analyses. **a)** Response to treatment, indicating improvement in MDMA symptoms per clinically significant change from baseline score (as defined by each study). Boxes represent the *RR* for each study, and the lines extending from the box represent the 95% confidence interval around each effect size, while the size of each box is proportional to its weight. The diamond at the bottom represents the pooled effect size (meta-analytic mean). The gray line at the bottom represents the prediction interval of the expected range of true effects in a new study. HK = Knapp-Hartung adjustment. **b)** Remission rates. Boxes represent the *RR* for each study, and the lines extending from the box represent the 95% confidence interval around each effect size, while the size of each box is proportional to its weight. The diamond at the bottom represents the pooled effect size (meta-analytic mean). The gray line at the bottom represents the prediction interval of the expected range of true effects in a new study. HK = Knapp-Hartung adjustment. pcb = placebo. **c)** Depression outcomes. Boxes represent the standardized mean difference (Hedges’ *g*) for each study, and the lines extending from the box represent the 95% confidence interval around each effect size, while the size of each box is proportional to its weight. The diamond at the bottom represents the pooled effect size (meta-analytic mean). The gray line at the bottom represents the prediction interval of the expected range of true effects in a new study. HK = Knapp-Hartung adjustment.

### Sensitivity analyses provide convergent results

We performed a series of sensitivity analyses that supported our primary results. First, our model using medium-dose arms in place of the high-dose arms for three-arm trials showed a significant and comparable effect size (**SI Fig. 2**; Hedges’ *g* = -0.75 [-1.19; -0.30], *p* = 0.008, *k* = 6, *n* = 234, *tau^2^* = 5.31 x 10^-6^ [0.00; 3.11], *I^2^* = 37.2% [0.0%; 75.0%]). Furthermore, we replicated our primary analyses using a fixed effect model on continuous outcomes (Hedges’ *g* = -0.71 [-0.98; -0.45], *p* < 0.001, *k* = 6, *n* = 242), response outcomes (*RR* = 1.44 [1.20; 1.74], *p* < 0.001, *k* = 5, *n* = 222), and remission outcomes (*RR* = 2.49 [1.52; 4.09], *p* < 0.001, *k* = 4, *n* = 210). Our Bayesian analysis revealed a Hedges’ *g* posterior distribution centered at -0.70 [-1.05; -0.36] (**SI Fig. 3**). Finally, our three-level CHE results were consistent across a range of within study correlation coefficients (**SI Fig. 4**).

### Overall low certainty GRADE rating

Evidence derived from randomized controlled trials begins with a high certainty rating that can be downgraded to moderate, low, or very low depending on assessments across four domains: risk of bias, inconsistency, indirectness, and imprecision. Here, we downgraded the certainty of evidence by two levels for indirectness. Functional unblinding, particularly in the two large phase 3 trials that used an inert placebo as comparator, presents risks for expectancy effects to drive larger between-group differences in outcomes. Further, the percentage of participants with prior MDMA use in the studies was high (mean: 39%). Taken together, these factors of indirectness may lead to smaller treatment effects in a general population under routine clinical practice and contribute to an overall GRADE rating of low certainty. No downgrades were given for risk of bias, inconsistency, or imprecision.

## Discussion

There is an urgent need for new therapies for PTSD (Imel et al., 2013; Sciarrino et al., 2020; Sripada et al., 2019). Current evidence, synthesized here, suggests that MDMA-assisted psychotherapy may have potential for treating PTSD. Compared to placebo, patients who received MDMA had significantly lower PTSD symptoms and higher rates of both response and remission. Furthermore, we found that a higher number of dosing sessions and higher cumulative MDMA dose was associated with larger improvements in PTSD symptoms. We present these results alongside an open-resource database, codebase, and dashboard - providing a public living data resource that will be regularly updated as new evidence emerges.

Preclinical and human studies suggest MDMA facilitates therapeutic mechanisms relevant to PTSD treatment. In rodents, MDMA (especially the *R*-entantiomer) promotes prosocial behaviors and fear extinction through serotonergic pathways (Curry et al., 2018; Young et al., 2017). In humans, MDMA shifts autobiographical recall toward more positive affect while preserving memory fidelity but reducing emotional salience (Carhart-Harris et al., 2014; Doss et al., 2018), as well as enhancing fear extinction learning (Maples-Keller et al., 2022; Vizeli et al., 2022). Neuroimaging in PTSD patients post-treatment shows altered activation in regions associated with fear response and autobiographical memory (S. P. Singleton et al., 2023). These findings suggest MDMA may support psychotherapy by reducing barriers to processing emotionally difficult content, enhancing the therapeutic alliance, and promoting memory reconsolidation and extinction (Feduccia & Mithoefer, 2018).

Our work aligns with and builds upon the existing literature in several important ways. First, while our results are consistent with prior meta-analyses, previous systematic reviews on the topic have quickly become outdated (Bahji et al., 2020, 2025; Brett & Bynum, 2025; Højlund et al., 2025; Hood et al., 2024; Hoskins et al., 2021; Illingworth et al., 2021; Kisely et al., 2023; Shahrour et al., 2024; Smith et al., 2022; Stanicic et al., 2025; Sze Jing Yong et al., 2025; Yang et al., 2024b; Žuljević et al., n.d., 2025). Second, in addition to our commitment to updating these results as a living review, our data and code are publicly accessible - maximizing transparency and reproducibility. To maximize transparency further, we also include detailed supplementary data and information, including a record of contacting study authors, a record of the amount of psychotherapy and psychological support provided in each study, the exact search terms used to find randomized controlled trials in each scientific database, and detailed extended methods.

Third, we also tested the robustness of primary findings through several sensitivity analyses, limiting the effects of analytic decisions on results.

Finally, our three-level CHE model and meta-regressions provide new results suggesting that MDMA’s effects may be amplified with an increased number of doses and a larger cumulative dose. Cumulative dose and the number of dosing sessions are both inherently intertwined, and thus our analyses are unable to distinguish between the two factors. Nonetheless, our analyses suggest that the largest between-group difference in PTSD scores occurs after the first dosing and integration sessions (Hedges’ *g* = -0.39), while additional dosing and integration sessions increase this difference by -0.20 SMD. However, the data included only span from one to three dosing sessions. This suggests longer trials may be useful to determine if further improvements may accumulate, and at what point additional dosing sessions no longer provide added clinically relevant benefits.

There remain substantial challenges in synthesizing evidence from currently available data (van Elk & Fried, 2023). Most MDMA for PTSD trials to date have been small (around 20 participants), with only two phase 3 trials recruiting closer to 100 participants (Mitchell et al., 2021, 2023). Furthermore, funding, design and implementation for all studies was guided by the same organization, the Multidisciplinary Association for Psychedelic Studies (MAPS) and their drug-development spin-off Resilient Pharmaceuticals (formerly Lykos Therapeutics) (Emerson et al., 2014), possibly contributing to the low heterogeneity observed in our models. However, there remains variability in the number of doses given to participants in the intervention and control groups, the timing of the primary endpoint, as well as the control methods (low-dose MDMA vs placebo). Furthermore, although none of the studies included in our database had a high risk of bias, our risk of bias assessments do not fully account for potential bias from functional unblinding or expectancy effects. Double-blinding in psychedelic-assisted psychotherapy has been the topic of extensive discourse in the field (Butler et al., 2022; Muthukumaraswamy et al., 2025; Schenberg, 2021; Szigeti & Heifets, 2024). The psychoactive and cardiovascular effects of MDMA make functional unblinding likely, and most studies that have formally assessed functional unblinding find that the majority of participants and/or blinded study staff correctly guess group assignments (Mitchell et al., 2023; Mithoefer et al., 2011, 2018; Oehen et al., 2013; Ot’alora G et al., 2018). Given the potential for functional unblinding, our large pooled effect size should be interpreted with caution.

Several limitations should be noted. Our meta-analysis included only a small number of studies (*k* = 6) that met eligibility criteria. MAPS and Resilient Pharmaceuticals/Lykos Therapeutics provided oversight of all of these studies, and the Phase 3 study data (Mitchell et al., 2021, 2023) was used in a new drug application to the FDA which was later denied (U.S. Food and Drug Administration, 2024). The FDA’s complete response letter cited three main concerns regarding the application. First, the failure to collect “positive” or “favorable” adverse events that may be relevant for abuse potential limited the assessment of the safety of MDMA therapy. We did not assess the safety of MDMA therapy here; this is the subject of a planned future analysis (S. P. Singleton et al., 2025). Second, evidence on the durability of the drug effect on outcomes was questionable. In our database, available endpoint data from longer durations was sparse, and longer duration studies are needed to determine the durability of MDMA’s effects beyond two months. Third, the possibility of selection bias during screening and the high rates of prior MDMA use in the study samples may have led to expectancy bias. As additional studies become available, meta-regression may be used to study this potential bias. Study participants were often carefully selected and may not be representative of the general population that might receive treatment if MDMA receives regulatory approval. As such, the data evaluated here may fail to generalize to a typical treatment-seeking population that includes high rates of both psychiatric, medical, and substance use co-morbidity (Lieberman et al., 2005; Miklowitz et al., 2007; Parikh et al., 2010; Stroup et al., 2006; Trivedi et al., 2006; Warden et al., 2007). Future studies with larger treatment groups and expanded inclusion criteria are essential to evaluate the generalizability of these findings. Lastly, it is worth noting that one high profile ethical violation occurred at a MAPS Phase 2 study site (Multidisciplinary Association for Psychedelic Studies, 2022). This site (*n* = 6; NCT01958593), along with one other small Phase 2 site (*n* = 8; NCT01689740), was not included in our database or analyses due to insufficient reporting.

The factors discussed above contributed to our low certainty GRADE rating for the evidence in this review. A low certainty rating indicates that the true effect of MDMA in ordinary clinical practice may be smaller than what has been observed in the literature so far. Thirteen phase 2 or 3 trials are currently active to evaluate MDMA for PTSD (*Search ClinicalTrials.Gov For*, n.d.). As new trials are published, our living systematic review, meta-analysis, and open science resource will continue to be updated to keep pace with the rapidly evolving evidence base.

## Conclusions

Together, synthesis of studies to date suggest that MDMA-assisted psychotherapy results in reductions in PTSD symptoms and increased response and remission in patients with PTSD. However, our GRADE rating for the certainty of this evidence is low. Additional, large controlled trials with rigorous methods are needed, as are studies examining varying study characteristics (e.g., dosing sessions, longer follow-up durations) and expanding to more representative populations. As more RCTs are published, we will regularly update our SYPRES website and dashboard in a reproducible and transparent manner. As part of our SYPRES initiative, we will also conduct a series of future meta-analyses on other psychedelic therapies, including LSD for anxiety and meta-analyses of safety outcomes (S. P. Singleton et al., 2025). This living systematic review and open science resource will provide a valuable and transparent resource for researchers, clinicians, policymakers, and the public.

## Article information

### Author contributions

BLS and SPS are joint first authors. JCS and TDS are joint senior authors.

Concept and design: All authors.

Acquisition, analysis, or interpretation of data: All authors.

Drafting of the manuscript: BLS, SPS, JCS, and TDS.

Critical revision of the manuscript for important intellectual content: All authors.

Statistical analysis: BLS, SPS, MJ, SNV, JCS, TDS.

Obtained funding: RHD and TDS.

Administrative, technical, or material support: PM and MH.

Supervision: JCS and TDS.

### Conflict of interest disclosures

Over the last three years, ECS has had grant funding to his institution from NIH; editing payments from: Wolters-Kluwer; compensation for serving on boards or consulting from: Eli Lilly and Company, Boehringer Ingelheim, Dimerx; medical devices supplied to his institution for his research: Masimo; and he has conducted medical-legal consultations. He also has served on the board of directors (unpaid) for a treatment program: Ashley Addiction Treatment. SMN is a co-investigator on a Usona Institute sponsored trial of psilocybin for Major Depressive Disorder. DBY has received research support from the NIH, the Gracias Family Foundation, the Heffter Research Institute, and philanthropic donors to the Johns Hopkins Center for Psychedelic and Consciousness Research. He co-directs the Hub at Oxford for Psychedelic Ethics (HOPE), which has received a small anonymous donation to support travel for underrepresented workshop participants; DBY does not personally receive financial support for this role. DBY has received a consulting fee from Soneira and educational speaking fees from the Integrative Psychiatry Institute. RHD, since 1 January 2021, has received research grants and contracts from the US FDA and the US NIH, and compensation for serving on advisory boards or consulting on clinical trial methods from Acadia, Akigai, Allay, AM-Pharma, Analgesic Solutions, Beckley, Biogen, Biosplice, Bsense, Cardialen, Chiesi, Clexio, Collegium, CombiGene, Confo, Contineum, Eccogene, Editas, Eli Lilly, Emmes, Endo, Epizon, Ethismos (equity), Exicure, GlaxoSmithKline, Glenmark, Gloriana, JucaBio, Kriya, Mainstay, Merck, Mind Medicine (also equity), NeuroBo, Noema, OliPass, Orion, Oxford Cannabinoid Technologies, Pfizer, Q-State, Regenacy (also equity), Rho, Salvia, Sangamo, Semnur, SIMR Biotech, Sinfonia, SK Biopharmaceuticals, Sparian, SPM Therapeutics, SPRIM Health, Tiefenbacher, Validae, Vertex, Viscera and WCG. All other authors have no conflicts to declare.

### Funding/support

This effort is supported by the Analgesic, Anesthetic, and Addiction Clinical Trial Translations, Innovations, Opportunities, and Networks and Pediatric Anesthesia Safety Initiative (ACTTION/PASI) public–private partnership with the US FDA. The views expressed in this article are those of the authors, and no endorsement by the FDA should be inferred. RHD is director and chair of the ACTTION management committee, and TDS obtained funding for this study via ACTTION.

### Additional contributions

We thank Jen Lege-Matsuura and Kristina McShea for their assistance with generating database search terms.

## Data Availability

The living database used for this analysis can be accessed through Metapsy (docs.metapsy.org/databases/ptsd-mdmactr/) or downloaded from our website, where code for all analyses is also openly available (sypres.io).

https://sypres.io

