## Supplementary Information for "A living systematic review, meta-analysis, and open data resource of trials of MDMA-assisted therapy for PTSD"

### Supplement 1

### Supplementary Figures

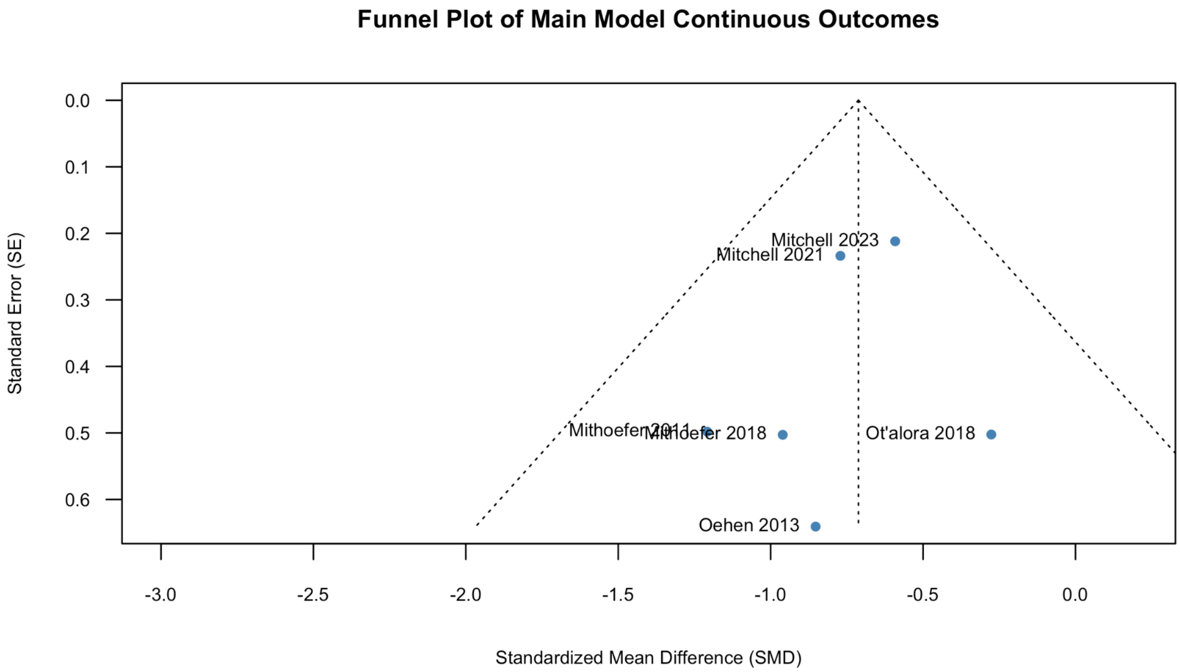

**SI Fig. 1**: Funnel plot of 6 effect sizes in the primary meta-analytic model.

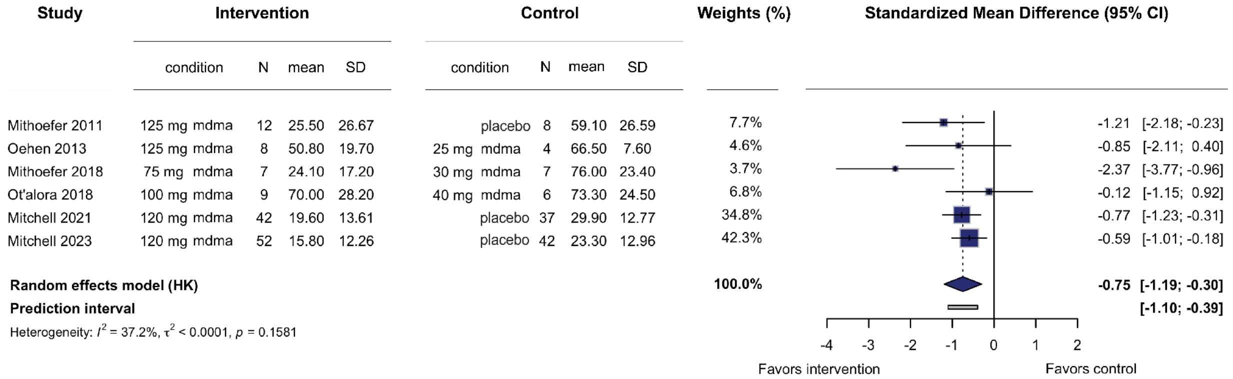

**SI Fig. 2: Meta-analysis on continuous outcomes using medium vs. low dose comparison in three-arm trials.** Two studies included in our database (Mithoefer 2018 and Ot'alora 2016) had 3 arms: a high dose of MDMA, a medium dose of MDMA, and a low dose. In our primary meta-analysis (see Fig 2), we compared the high dose of MDMA to the low dose; in this figure, we instead used the medium dose of MDMA compared to the same low dose in those two studies. Boxes represent the standardized mean difference (Hedges' *g*) for each study, and the lines extending from the box represent the 95% confidence interval around each effect size, while the size of each box is proportional to its weight. The diamond at the bottom represents the pooled effect size (meta-analytic mean). The gray line at the bottom represents the prediction interval of the expected range of true effects in a new study. HK = Knapp-Hartung adjustment.

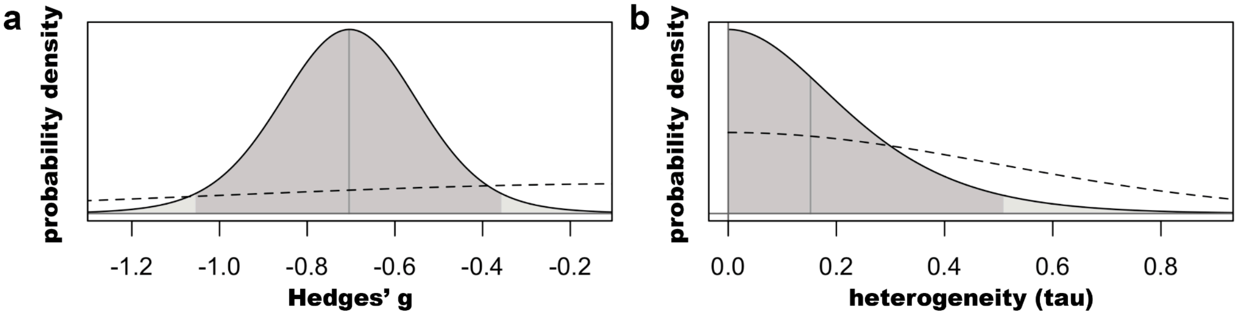

**SI Fig. 3:** Prior (dashed) and posterior (solid) distributions for (**a**) the pooled effect size (Hedges' *g*) and (**b**) the heterogeneity estimate (*tau*). Dark shading represents the 95% probability distribution.

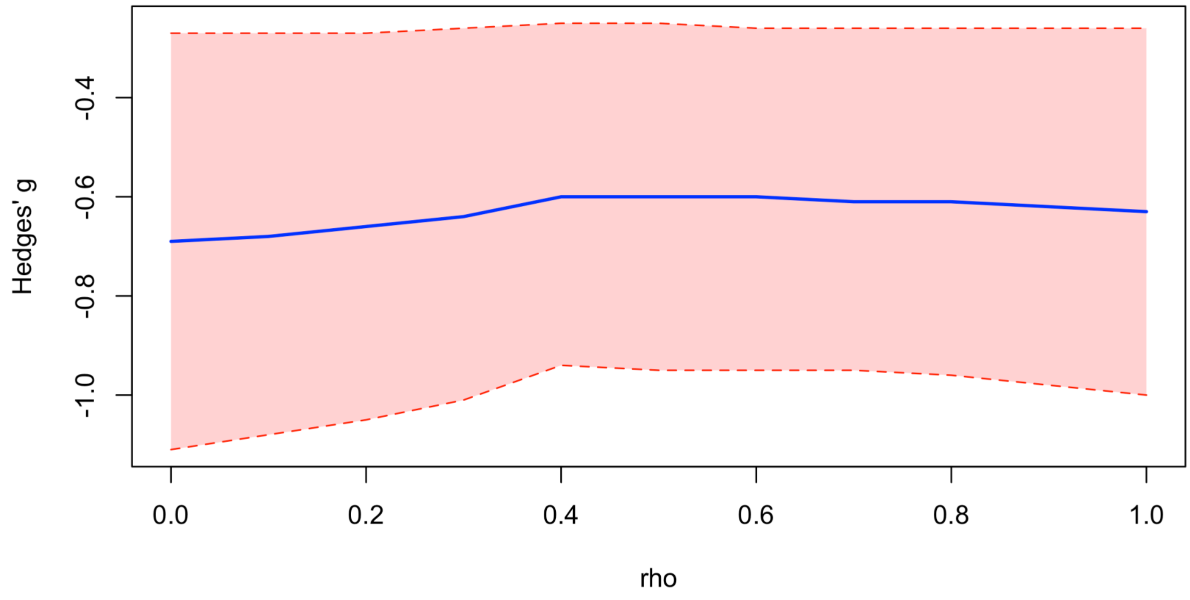

**SI Fig. 4:** Hedges' *g* as a function of *rho* ($\rho$), the within-study correlation coefficient used in the three-level CHE model. The blue line represents the estimated pooled effect, and the shaded red region represents the confidence interval.

### Supplementary Methods

#### Search and selection

We searched PubMed, Embase, PsycInfo, Web of Science, and Scopus. Additionally, we searched reference lists of prior systematic reviews identified from our primary search. Our search syntax combined MeSH and text terms under the PICO (Population, Intervention, Comparator, Outcome) framework to identify randomized controlled trials on the effects of MDMA on PTSD (see *Search Terms* for a complete list of search syntax for each database). The search syntax was developed in collaboration with research librarians with expertise in systematic reviews. The initial database searches occurred on April 20. The final search was conducted on November 25, 2025. The publication date range for included studies was from the database start range to the final search date. Updated searches will be conducted at least annually given the living nature of this systematic review, with updated results published on our SYPRES website.

Search results were saved and loaded into Covidence, a web-based collaboration software platform that streamlines the production of systematic and other literature reviews. Covidence automatically filtered out duplicates and auto-marked non-RCTs as ineligible. Two team members (SPS & BLS) performed screening of articles based on predefined exclusion and inclusion criteria (see *Eligibility Criteria* in the main text). In screening, we evaluated the title and abstract of all references returned by our search. If both reviewers voted to include the reference in full text review, then it was moved forward to the next phase of evaluation. Conflicts were resolved via a consensus discussion before the next phase. In the next phase, both reviewers independently evaluated the full text of the article. Disagreements on either inclusion or exclusion (or on the reason for exclusion) were resolved via a consensus discussion or discussion with a third reviewer (TDS or JCS). Reasons for excluding a report were determined on a 5-level hierarchical basis. First, if the report was not an RCT or presented in a language other than English, it was excluded. Second, if neither of these applied, but the report represented a published protocol of an upcoming RCT or initial results from an incomplete RCT, it was excluded. Third, if neither of the first two levels applied, but if the report was a preprint or conference abstract of a study where future peer-reviewed results were anticipated but not yet published, it was excluded. Fourth, if the report did not present any mood outcomes or if it was the wrong patient population (e.g. healthy individuals or pediatric patients) then it was excluded. Fifth and finally, if none of the previous levels applied, then any of the following reasons could qualify as an exclusion reason: the report was a conference abstract of a study where the full-text was later published, or a secondary report of an already included original study, or the study report included insufficient data for a meta-analysis. If none of these reasons applied, the report was included. If a secondary report contained information possibly relevant to current or future versions of our database, we included it and merged it with the parent study. See *Excluded Reports* for the full list of reports excluded during full-text review and their reasons. The two reviewers agreed on 96% and 100% of title/abstract screening and full-text review decisions, respectively. Complete details of the study selection process including reasons for exclusion are provided in **Figure 1**.

#### Data extraction

Data was independently extracted from included studies by two team members (BLS & AL or SPS) using Covidence. Resources used to extract the data included the primary paper, supplemental data, clinical trial registries, and protocol details. Follow-up papers published on the same clinical trial that either added additional time points or more mood-related outcomes to the original paper were grouped with the original paper and extracted as one study to avoid duplication.

Extracted data included identification information, such as author details, setting, and time frame of clinical trial; methods information, such as study design, randomization and recruitment methods, blinding, assessment and therapy details, outcome details, the primary time-point and outcome, and statistical analyses used; population information, such as inclusion and exclusion criteria, withdrawal details, and baseline characteristics; intervention information, such as allocation to groups, dosage information, and frequency of drug administration; and outcomes and results information. For outcomes and results, we extracted all relevant PTSD and mood outcomes at every timepoint provided in the paper. We extracted both dichotomous and continuous outcome measures in the format provided by the paper or supplementary data, prioritizing raw data over effect sizes. The dichotomous outcomes of response and remission were defined internally within each study and not by the authors of this review.

If information pertinent to the meta-analysis were not available, authors were contacted via e-mail. If desired data were available only in figures and not reported in the article text or successfully obtained via author contact, we used the WebPlotDigitizer tool[2] to extract results data.

Disagreements in data extraction were resolved via consensus discussion or by a third reviewer (SPS). For WebPlotDigitizer data, one team member (SPS) used the tool to extract results, while another team member (AL) checked the results. In case of disagreement, the second reviewer replicated the data extraction using the WebPlotDigitizer tool, and remaining disagreements were resolved by a consensus discussion.

#### Risk of bias assessment

We assessed study bias using Cochrane's Risk of Bias (RoB) 2.0 tool[3,4], which is the standard approach for bias assessment in randomized controlled trials. We focused our assessment specifically on primary outcome variables. In studies with multiple primary outcomes, we evaluated the outcome selected for our primary meta-analysis (see *Selection of Effect Sizes* section in supplement). Furthermore, we limited our bias assessment to pre-crossover study periods, as our analyses here were restricted to pre-crossover data.

The RoB 2.0 tool examines five potential bias sources: randomization procedures, deviations from the intended protocol, missing outcome data, outcome measurement, and selective reporting. Each domain contains several questions rated on a four-level scale ranging from 'yes' to 'no', in addition to an option for insufficient information. Domain-level and overall study bias ratings are classified as low, medium, or high according to a predetermined algorithm[4]. While this algorithm guides assessment, evaluators may justifiably override automated bias determinations when specific concerns warrant greater or lesser emphasis than the algorithm suggests. Two team members (SPS & BLS) independently conducted the bias assessment. Through collaborative discussion and re-examination, at times with additional team members (SMN & JCS), consensus was ultimately achieved. **Table 2** presents the final assessments; Supplement 2 details individual items.

We used the handbook associated with the RoB 2.0 Tool to interpret the signaling questions, but some of the questions had particular nuances in the context of psychedelic-assisted therapy, which were discussed by team members before conducting RoB assessments to ensure consistent interpretations. See *Risk of Bias Assessment Standard Operating Procedure* for the full risk of bias assessment procedure that was used in conjunction with the Cochrane tool guidelines and handbooks to answer signaling questions.

An important consideration of psychedelic-assisted psychotherapy trials is the high potential for functional unblinding due to the unique psychoactive and cardiovascular effects of MDMA. We took this into account when answering risk of bias questions in both Domain 2, focusing on protocol deviations, and Domain 4, focusing on the outcome measurement. Signaling questions 2.1 and 2.2 ask respectively if participants and carers or people delivering the intervention were aware of participants' assigned intervention during the trial. Although most of the studies in our meta-analyses were methodologically double-blind studies, due to the psychoactive and cardiovascular effects in the MDMA group, we answered 'PY' to this question in most cases. If functional unblinding was measured in either participants and/or study staff, we answered 'Y' if the results suggested unblinding and 'N' if the results suggested successful blinding. Oehen 2013 was the only study where the authors concluded blinding was successful. Note that while our answer of 'Y' for studies that actually assessed functional unblinding indicates an increased confidence in the answer to the signaling question, in the risk of bias algorithm responses of 'PY' and 'Y' are weighted equally.

In domain 4, signaling question 4.3 asked if outcome assessors were aware of the intervention received by study participants. In this case, the relevance of functional unblinding depended on who the outcome assessors were. For self-report questionnaires, the outcome assessor was the participant. For clinician-administered questionnaires, outcome assessors were either blinded raters (such as independent study staff or third party raters), or unblinded raters (such as a non-independent study staff who were involved in the administration of therapy or other forms of patient contact). Following the same logic used when answering signaling question 2.1 and 2.2, we answered 4.3 as follows. Self-report questionnaires and unblinded (functionally or otherwise) clinician raters received 'PY' or 'Y', and clinician raters who were blinded independent study staff or third-party raters received 'PN' or 'N'. Answering 'PN' or 'N' on 4.3 ends the algorithm and signals a low risk of bias for domain 4. Thus, studies that used blinded independent study-staff or third-party raters to administer clinician assessments received a low risk of bias for this domain. If any other answer is given for 4.3, two additional questions follow. Specifically, 4.4 asks if knowledge of the intervention *could* influence the assessment, to which we always answered 'Y', and 4.5 asks if knowledge of the intervention *is likely* to influence the assessment. Here, we considered self-report data to have less risk of bias than explicitly unblinded (open-label) non-independent study-staff rating clinician-administered scales. A non-independent study staff potentially had more bias or stake in the "success" of treatment, potentially contributing to bias in the way they surveyed the outcome measure, requiring an answer of 'PY' for 4.5. However, there is evidence that self-reports actually have low risk of bias and do not over-inflate the effects of the intervention treatment [5], justifying a response of 'PN' for 4.5. For clinician-administered outcomes by other non-independent study staff, a judgement call must be made based on their level of patient contact, potential for functional unblinding, and overall involvement in the study. Domain 4 assessment is summarized in **SI Table 1** below:

| Assessor | Domain 4 Result |
|---|---|
| Blinded third-party independent rater | Low |
| Blinded independent study staff | Low |
| Potentially functionally unblinded non-independent study staff | Some concerns or High[a] |
| Explicitly unblinded (open-label) non-independent study staff | High |
| Self | Some concerns |

**SI Table 1:** Summary of the relationship between the identity of the assessor and risk of bias rating on domain 4. [a]May be upgraded to 'High' depending on the rater's level of patient contact, potential for functional unblinding, and overall involvement in the study.

#### Selection of effect sizes

For all models except the three-level correlated and hierarchical effects (CHE) model (see below), only one effect size was used per study. In most cases, this was the Hedges' *g* calculated at that study's primary endpoint, using their own reported primary outcome PTSD instrument. For all analyses, only data from pre-crossover timepoints were used. For studies that did not report a primary PTSD instrument, or reported multiple, we identified one to serve as primary (**SI Table 2**). In these cases, we first prioritized clinician-administered instruments (i.e., CAPS), when available. For studies that did not report a primary endpoint, or reported multiple primary endpoints, we selected the last available time point that was within four weeks of the average time across other included studies and prior to any crossover event, open-label extension, or follow-up period (**SI Table 3**). For the five studies which reported dichotomous outcomes, we used effect sizes (risk ratios) for the same instrument and timepoint as was used in the primary continuous analysis. For our three-level CHE model on continuous outcomes, we used any effect size extracted from a study that was derived from the selected primary instrument during any time point in the post-dosing study period prior to any crossover event or follow-up period (**SI Table 4**). See **SI Table 5** for the full list of outcomes and timepoints extracted in each study.

| Study | Reported Primary Instrument(s) | Selected Instrument |
|---|---|---|
| Mithoefer 2011 | CAPS-IV | CAPS-IV |
| Oehen 2013 | CAPS-IV; PDS | CAPS-IV |
| Mithoefer 2018 | CAPS-IV | CAPS-IV |
| Ot'alora 2018 | CAPS-IV | CAPS-IV |
| Mitchell 2021 | CAPS-V | CAPS-V |
| Mitchell 2023 | CAPS-V | CAPS-V |

**SI Table 2**: Primary instruments reported by each study along with the instrument used for our analyses. CAPS-IV = Clinician-Administered PTSD Scale (v4); CAPS-V = Clinician-Administered PTSD Scale (v5); PDS = Posttraumatic Diagnostic Scale (PDS)

| Study | Reported Primary Endpoint(s) | Selected Endpoint |
|---|---|---|
| Mithoefer 2011 | 3-5 days after first experimental session; 3-5 days after second experimental session; 2 months after second experimental session | 2 months after second experimental session |
| Oehen 2013 | 3 weeks after third dose; 2 months*; 6 months* | 3 weeks after third dose |
| Mithoefer 2018 | Month 1 after second experimental session | Month 1 after second experimental session |
| Ot'alora 2018 | 1 month after two sessions | 1 month after two sessions |
| Mitchell 2021 | Week 18 (after third dose) | Week 18 (after third dose) |
| Mitchell 2023 | Week 18 (after third dose) | Week 18 (after third dose) |

**SI Table 3**: Primary endpoints reported by each study along with the endpoint selected for our analyses. *These timepoints occurred after an open-label crossover extension period.

| Study | Comparators (arm 1 vs. arm 2) | Instrument | Timepoint | N arm 1 | Mean (SD) arm 1 | N arm 2 | Mean (SD) arm 2 |
|---|---|---|---|---|---|---|---|
| Mithoefer 2011 | 125 mg MDMA vs. 125 mg placebo | CAPS-IV | 3-5 days after first session | 12 | 37.8 (29.1) | 8 | 74.1 (29.1) |
| | | | 3-5 days after second session | 12 | 29.3 (22.5) | 8 | 66.8 (22.6) |
| | | | 2 months after second session | 12 | 25.5 (26.7) | 8 | 59.1 (26.6) |
| Oehen 2013 | 125 mg MDMA vs. 25 mg MDMA | CAPS-IV | 3 weeks after second dose | 8 | 63.0 (17.8) | 4 | 60.0 (6.8) |
| | | | 3 weeks after third dose | 8 | 50.8 (19.7) | 4 | 66.5 (7.6) |
| Mithoefer 2018 | 125 mg MDMA vs. 30 mg MDMA | CAPS-IV | 1 month after second experimental session | 12 | 45.3 (33.8) | 7 | 76.0 (23.4) |
| | 75 mg MDMA vs. 30 mg MDMA | | | 7 | 24.1 (17.2) | 7 | 76.0 (23.4) |
| Ot'alora 2018 | 125 mg MDMA vs. 40 mg MDMA | CAPS-IV | 1 month after two sessions | 12 | 64.3 (33.6) | 6 | 73.3 (24.5) |
| | 100 mg MDMA vs. 40 mg MDMA | | | 9 | 70.0 (28.2) | 6 | 73.3 (24.5) |
| Mitchell 2021 | 80/120 mg MDMA vs. 80/120 mg placebo | CAPS-V | Week 4 (3 weeks after dose 1) | 46 | 33.8 (12.2) | 43 | 37.0 (10.5) |
| | | | Week 8 (3 weeks after dose 2) | 42 | 26.3 (12.3) | 39 | 33.5 (12.5) |
| | | | Week 18 (9 weeks after dose 3) | 42 | 19.6 (13.6) | 37 | 29.9 (12.8) |
| Mitchell 2023 | 80/120 mg MDMA vs. 80/120 mg placebo | CAPS-V | Week 4 (3 weeks after dose 1) | 53 | 28.1 (13.8) | 50 | 31.5 (9.2) |
| | | | Week 8 (3 weeks after dose 2) | 53 | 20.8 (13.1) | 44 | 27.7 (11.9) |
| | | | Week 18 (9 weeks after dose 3) | 52 | 15.8 (12.3) | 42 | 23.3 (13.0) |

**SI Table 4**: List of the raw data for the 15 effect sizes selected for the three-level CHE model. All timepoints are listed as reported by the study text with additional context added in parentheses for the Mitchell 2021 and Mitchell 2023 studies. The Mitchell 2021 and Mitchell 2023 studies escalated the dose of MDMA and placebo from 80 mg for the first session to 120 mg for the second and third sessions. All studies offered supplemental half-doses partway through the dosing sessions.

### Database description

Our publicly released database contains 61 total effect sizes generated from 6 studies (**SI Table 5**). These effect sizes encompass all PTSD outcomes and all timepoints reported by arm in each study. Effect sizes are generated from five different outcome types (**SI Table 6**). Secondary depression outcomes are not a part of the database, however the raw data used for this analysis can be found in **Figure 4**. Full, up-to-date documentation on the database, including descriptions of all variables, can be found here: https://docs.metapsy.org/databases/ptsd-mdmactr/.

| Study | Effect sizes (N) | Instruments | Timepoints |
|---|---|---|---|
| Mitchell 2021 | 9 | CAPS-V | Week 4, Week 8, Week 18 |
| Mitchell 2023 | 10 | CAPS-V | Week 4, Week 8, Week 18 |
| Mithoefer 2018 | 18 | CAPS-IV | Month 1 after second experimental session, Month 12 |
| Oehen 2013 | 7 | CAPS-IV, PDS | 3 weeks after second dose, 3 weeks after third dose |
| Mithoefer 2011 | 5 | CAPS-IV, IES-R | 3-5 days after first experimental session, 3-5 days after second experimental session, 2 months after second experimental session |
| Ot’alora 2018 | 12 | CAPS-IV | 1 month after 2 sessions |

**SI Table 5**: List of 61 effect sizes compiled in the full database released through Metapsy. Timepoints here are verbatim references to the text used in the study reports.

| Outcome type | Effect size | Inputs |
|---|---|---|
| msd | Hedges’ *g* | 1. Endpoint N’s<br>2. Endpoint means<br>3. Endpoint standard deviations |
| change | Hedges’ *g* | 1. Endpoint N’s<br>2. Endpoint means calculated by adding change score means to baseline means<br>3. Change score standard deviations[a] |
| response | Log Risk Ratio | 1. N responders at endpoint |

| | | |
|---|---|---|
| | | 2. N participants at endpoint |
| remission | Log Risk Ratio | 1. N remitters at endpoint<br>2. N participants at endpoint |

**SI Table 6**: Details on each outcome type, denoted by the variable `outcome_type` in the database. [a]This approach assumes a correlation coefficient of 0.5 between baseline and endpoint measurements. At this value, the standard deviations for change score and endpoint means are equal.

### Search Terms

#### Pubmed

**#1:** '("PTSD"[Title/Abstract] OR "post-traumatic stress"[Title/Abstract] OR "posttraumatic stress*"[Title/Abstract] OR "stress disorders, post-traumatic"[MeSH Terms] )
**#2:** '("psychedelic"[Title/Abstract] OR "MDMA"[Title/Abstract] OR “methylenedioxymethamphetamine”[Title/Abstract] OR "N-Methyl-3,4-methylenedioxyamphetamine"[MeSH Terms] OR "ecstasy"[Title/Abstract] OR "molly"[Title/Abstract]) OR “midomafetamine”[Title/Abstract]
**#3:** '("Randomized Controlled Trial"[Publication Type] OR "Controlled Clinical Trial"[Publication Type] OR "Pragmatic Clinical Trial"[Publication Type] OR "Equivalence Trial"[Publication Type] OR "clinical trial, phase iii"[Publication Type] OR "Randomized Controlled Trials as Topic"[MeSH Terms] OR "Controlled Clinical Trials as Topic"[MeSH Terms] OR "Random Allocation"[MeSH Terms] OR "Double-Blind Method"[MeSH Terms] OR "Single-Blind Method"[MeSH Terms] OR "placebos"[MeSH Terms:noexp] OR "Control Groups"[MeSH Terms] OR ("random"[Title/Abstract] OR "sham"[Title/Abstract] OR "placebo"[Title/Abstract]) OR (("singl*"[Title/Abstract] OR "doubl*"[Title/Abstract]) AND ("blind*"[Title/Abstract] OR "dumm*"[Title/Abstract] OR "mask*"[Title/Abstract])) OR (("tripl*"[Title/Abstract] OR "trebl*"[Title/Abstract]) AND ("blind*"[Title/Abstract] OR "dumm*"[Title/Abstract] OR "mask*"[Title/Abstract])) OR ("control*"[Title/Abstract] AND ("study"[Title/Abstract] OR "studies"[Title/Abstract] OR "trial*"[Title/Abstract] OR "group*"[Title/Abstract]))))
**#4: #1 AND #2 AND #3**

#### Embase

**#1:** '('ptsd':ti,ab OR 'post-traumatic stress':ti,ab OR 'posttraumatic stress*':ti,ab OR 'posttraumatic stress disorder'/exp)
**#2:** '('psychedelic':ti,ab OR 'mdma':ti,ab OR 'methylenedioxymethamphetamine':ti,ab OR 'midomafetamine'/exp OR 'ecstasy':ti,ab OR 'molly':ti,ab OR 'midomafetamine':ti,ab)

**#3:** '('randomized controlled trial':it OR 'controlled clinical trial':it OR 'pragmatic clinical trial':it OR 'equivalence trial':it OR 'clinical trial, phase iii':it OR 'randomized controlled trial (topic)'/exp OR 'controlled clinical trial (topic)'/exp OR 'randomization'/exp OR 'double blind procedure'/exp OR 'single blind procedure'/exp OR 'placebo'/de OR 'control group'/exp OR ('random':ti,ab OR 'sham':ti,ab OR 'placebo':ti,ab) OR (('singl*':ti,ab OR 'doubl*':ti,ab) AND ('blind*':ti,ab OR 'dumm*':ti,ab OR 'mask*':ti,ab)) OR (('tripl*':ti,ab OR 'trebl*':ti,ab) AND ('blind*':ti,ab OR 'dumm*':ti,ab OR 'mask*':ti,ab)) OR ('control*':ti,ab AND ('study':ti,ab OR 'studies':ti,ab OR 'trial*':ti,ab OR 'group*':ti,ab)))
**#4: #1 AND #2 AND #3**

#### PsycInfo

**S1:** (tiab("PTSD" OR "post-traumatic stress" OR "posttraumatic stress*") OR MAINSUBJECT.EXACT.EXPLODE("Posttraumatic Stress Disorder"))
**S2:** (tiab("psychedelic" OR "MDMA" OR "methylenedioxymethamphetamine" OR "ecstasy" OR "molly" OR "midomafetamine") OR MAINSUBJECT.EXACT.EXPLODE("Methylenedioxymethamphetamine"))
**S3:** (MAINSUBJECT.EXACT.EXPLODE("Randomized Controlled Trials" OR "Placebo" OR "Experiment Controls") OR (TIAB("random" OR "sham" OR "placebo" OR "Pragmatic Clinical Trial" OR "Equivalence Trial" OR "clinical trial, phase iii" OR "Random Allocation" OR "Double-Blind Method" OR "Single-Blind Method")) OR (TIAB("singl*" OR "doubl*") AND ("blind*" OR "dumm*" OR "mask*")) OR (TIAB("tripl*" OR "trebl*") AND ("blind*" OR "dumm*" OR "mask*")) OR TIAB("control*" AND ("study" OR "studies" OR "trial*" OR "group*")))
**#4: [S1] AND [S2] AND [S3]**

#### Web of Science

**#1:** ((TI=PTSD OR AB=PTSD) OR (TI="post-traumatic stress" OR AB="post-traumatic stress") OR (TI="posttraumatic stress*" OR AB="posttraumatic stress*") OR TS="stress disorders, post traumatic")
**#2:** ((TI=psychedelic OR AB=psychedelic) OR (TI=MDMA OR AB=MDMA) OR (TI=methylenedioxymethamphetamine OR AB=methylenedioxymethamphetamine) OR TS="n methyl 3,4 methylenedioxyamphetamine" OR (TI=ecstasy OR AB=ecstasy) OR (TI=molly OR AB=molly) OR (TI=midomafetamine OR AB=midomafetamine))
**#3:** (TS="Randomized Controlled Trial" OR TS="Controlled Clinical Trial" OR TS="Pragmatic Clinical Trial" OR TS="Equivalence Trial" OR TS="clinical trial, phase iii" OR TS="Randomized Controlled Trials as Topic" OR TS="Controlled Clinical Trials as Topic" OR TS="Random Allocation" OR TS="Double-Blind Method" OR TS="Single-Blind Method" OR TS=placebos OR TS="Control Groups" OR ((TI=random OR AB=random) OR (TI=sham OR AB=sham) OR (TI=placebo OR AB=placebo)) OR (((TI=singl* OR AB=singl*) OR (TI=doubl* OR AB=doubl*)) AND ((TI=blind* OR AB=blind*) OR (TI=dumm* OR AB=dumm*) OR (TI=mask* OR AB=mask*))) OR (((TI=tripl* OR AB=tripl*) OR (TI=trebl* OR AB=trebl*)) AND ((TI=blind* OR

AB=blind*) OR (TI=dumm* OR AB=dumm*) OR (TI=mask* OR AB=mask*))) OR ((TI=control* OR AB=control*) AND ((TI=study OR AB=study) OR (TI=studies OR AB=studies) OR (TI=trial* OR AB=trial*) OR (TI=group* OR AB=group*))))
**#4: #1 AND #2 AND #3**

#### Scopus

**#1:** ( TITLE-ABS ( "PTSD" OR "post-traumatic stress" OR "posttraumatic stress*" ) OR INDEXTERMS ( "stress disorders, post traumatic" ) )
**#2:** ((TITLE-ABS(psychedelic OR MDMA OR methylenedioxymethamphetamine OR ecstasy OR molly OR midomafetamine)) OR INDEXTERMS("n methyl 3,4 methylenedioxyamphetamine"))
**#3:** (DOCTYPE("Randomized Controlled Trial" OR "Controlled Clinical Trial" OR "Pragmatic Clinical Trial" OR "Equivalence Trial" OR "clinical trial, phase iii") OR INDEXTERMS("Randomized Controlled Trials as Topic" OR "Controlled Clinical Trials as Topic" OR "Random Allocation" OR "Double-Blind Method" OR "Single-Blind Method" OR placebos OR "Control Groups") OR TITLE-ABS(random OR sham OR placebo) OR ((TITLE-ABS(singl* OR doubl*)) AND (TITLE-ABS(blind* OR dumm* OR mask*))) OR ((TITLE-ABS(tripl* OR trebl*)) AND (TITLE-ABS(blind* OR dumm* OR mask*))) OR (TITLE-ABS(control*) AND (TITLE-ABS(study OR studies OR trial* OR group*))))
**#4: #1 AND #2 AND #3**

### Psychotherapy or Psychological Support

| Study | Psychotherapy Sessions and Frequency |
|---|---|
| Mithoefer 2011 | 2 sessions before dosing<br>1 integration session the morning after the 1st dose<br>3 weekly integration sessions<br>1 integration session the morning after the 2nd dose<br>3 weekly integration sessions<br>1 final integration session 2 months after the 2nd dose |
| Oehen 2013 | 2 preparatory sessions preceding the first MDMA session<br>3 MDMA-assisted sessions each approximately 8 hours<br>1 non-drug psychotherapy session the morning after each MDMA |

| | |
|---|---|
| | experience, followed by two integration sessions one week apart<br>1 telephone call following each MDMA-assisted session on a daily basis for 1 week<br>Subjects each received a total of 12 non-drug psychotherapy sessions. Additional sessions in case of excessive distress were limited to two after each MDMA session. |
| Mithoefer 2018 | 3 preparatory sessions before 1st dose<br>1 session during each dose (2 doses total)<br>1 integration session the morning after each dose<br>2 weekly sessions after dosing |
| Ot'aloraG 2018 | 3 90-minute preparatory sessions before MDMA sessions<br>3 integrative sessions after MDMA sessions (the first of the three integrative sessions was conducted on the morning following each experimental session)<br>Daily 15-60 minute telephone contact for 7 days following each experimental session<br>2 integrative sessions in between the two experimental sessions |
| Mitchell 2021 | 3 90-minute preparatory sessions<br>3 weekly 90-minute integration sessions after each dosing session (total of 3 doses) |
| Mitchell 2023 | 3 90-minute preparation sessions<br>3 90-minute integration sessions following each dose (the first integration session took place on the morning after the experimental session and was followed by phone check-ins every other day for 2 weeks. The second and third integration sessions occurred over 3-4 weeks following the first integration session). |

**SI Table 7**: Psychotherapy or psychological support sessions and their frequency for each study.

### Excluded Reports

| **Excluded Study Title:** | **Excluded Study Author and Year:** | **Reason for Exclusion:** |
|---|---|---|
| Self-compassion mediates treatment effects in MDMA-assisted therapy for posttraumatic stress disorder. | Agin-Liebes 2025[6] | Re-analysis/Secondary report |
| Incorporating MDMA as an adjunct in emotionally focused couples therapy with clients impacted by trauma or PTSD | Almond 2019[7] | Not an RCT |

| | | |
|---|---|---|
| Oxytocin and the Role of Fluid Restriction in MDMA-Induced Hyponatremia: A Secondary Analysis of 4 Randomized Clinical Trials. | Atila 2024[8] | Wrong patient population (e.g., healthy, pediatric) |
| Perceived Benefits of MDMA-Assisted Psychotherapy beyond Symptom Reduction: Qualitative Follow-up Study of a Clinical Trial for Individuals with Treatment-Resistant PTSD | Barone 2019[9] | Re-analysis/Secondary report |
| MDMA-assisted therapy significantly reduces eating disorder symptoms in a randomized placebo-controlled trial of adults with severe PTSD | Brewerton 2022[10] | Re-analysis/Secondary report |
| MDMA-assisted psychotherapy using low doses in a small sample of women with chronic posttraumatic stress disorder | Bouso 2008[1] | Insufficient data |
| Discontinuation of medications classified as reuptake inhibitors affects treatment response of MDMA-assisted psychotherapy. | Feduccia 2021[11] | Re-analysis/Secondary report |
| Posttraumatic Growth After MDMA-Assisted Psychotherapy for Posttraumatic Stress Disorder. | Gorman 2020[12] | Re-analysis/Secondary report |
| Long-term follow-up outcomes of MDMA-assisted psychotherapy for treatment of PTSD: a longitudinal pooled analysis of six phase 2 trials. | Jerome 2020[13] | Re-analysis/Secondary report |
| Pilot study suggests DNA methylation of the glucocorticoid receptor gene (NR3C1) is associated with MDMA-assisted therapy treatment response for severe PTSD. | Lewis 2023[14] | Re-analysis/Secondary report |
| MDMA-Therapy for Trauma Related Disorders | Mitchell 2023[15] | Conference abstract - full text already included |
| MDMA-assisted psychotherapy for treatment of PTSD: study design and rationale for phase 3 trials based on pooled analysis of six phase 2 randomized controlled trials. | Mithoefer 2019[16] | Re-analysis/Secondary report |
| The effects of MDMA-assisted therapy on alcohol and substance use in a phase 3 trial for treatment of severe PTSD | Nicholas 2022[17] | Re-analysis/Secondary report |
| Sleep Quality Improvements After | Ponte 2021[18] | Re-analysis/Secondary |

| | | |
|---|---|---|
| MDMA-Assisted Psychotherapy for the Treatment of Posttraumatic Stress Disorder. | | report |
| Mdma-assisted psychotherapy for the treatment of ptsd | Reiff 2021[19] | Not an RCT |
| Altered brain activity and functional connectivity after MDMA-assisted therapy for post-traumatic stress disorder | Singleton 2023[20] | Re-analysis/Secondary report |
| Efficacy and safety results from the first pivotal phase 3 randomized controlled trial of mdma-assisted psychotherapy for treatment of severe chronic PTSD | van der Kolk 2021[21] | Conference abstract - full text already included |
| Effects of MDMA-assisted therapy for PTSD on self-experience. | van der Kolk 2024[22] | Re-analysis/Secondary report |
| A Randomized, Double-Blind, Placebo Controlled Phase 3 Study Assessing Efficacy and Safety of MDMA-Assisted Therapy for the Treatment of Severe PTSD | Yazar-Klosinski 2021[23] | Conference abstract - full text already included |
| Preliminary evidence for the importance of therapeutic alliance in MDMA-assisted psychotherapy for posttraumatic stress disorder. | Zeifman 2024[24] | Re-analysis/Secondary report |

**SI Table 8**: List of reports excluded during full-text review. Excluded secondary reports also appear in the next table.

### Merged and Excluded Secondary Reports

| **Primary study:** | **Merged report:** | **What does the merged report(s) add to the database?** | **Additional secondary reports that were excluded in full-text review:** |
|---|---|---|---|
| Mithoefer 2011 | Mithoefer 2011[25]<br>Mithoefer 2013[26] | Correction<br>Long-term follow-up[a] | Zeifman 2024[24] |
| Mithoefer 2018 | N/A | N/A | Barone 2019[9]<br>Singleton 2023[20] |
| Mitchell 2021 | N/A | N/A | Brewerton 2022[10]<br>Nicholas 2022[17] |

| | | | |
|---|---|---|---|
| | | | Lewis 2023[14]<br>van der Kolk 2024[22] |
| Mitchell 2023 | N/A | N/A | Agin-Liebes 2025[6] |

**SI Table 9**: Record of secondary reports that were either merged with the parent study for extraction or excluded in full-text screening. [a]Long-term follow-up data was after unblinding, so data from this paper was not extracted in our database.

| **Secondary report on pooled studies:** | **Primary studies (MAPS number, clinicaltrials.gov identifier):** |
|---|---|
| Feduccia 2021[11]* | Mithoefer 2018[27] (MP-8; NCT01211405)<br>Ot'alora 2018[28] (MP-12; NCT01793610)<br>Unpublished (MP-4; NCT01958593)<br>Unpublished (MP-9; NCT01689740) |
| Gorman 2020[12*] | Mithoefer 2018[27] (MP-8; NCT01211405)<br>O'talora G 2018[28] (MP-12; NCT01793610)<br>Unpublished (MP4; NCT01958593) |
| Jerome 2020[13]* | Mithoefer 2011[29] (MP-1; NCT00090064)<br>Oehen 2013[30] (MP-2; NCT00353938)<br>Mithoefer 2018[27] (MP-8; NCT01211405)<br>Ot'alora 2018[28] (MP-12; NCT01793610)<br>Unpublished (MP-4; NCT01958593) |

| | |
|---|---|
| | Unpublished (MP-9; NCT01689740) |
| Mithoefer 2019[16]* | Mithoefer 2011[29] (MP-1; NCT03537014)<br>Oehen 2013[30] (MP-2; NCT04077437)<br>Mithoefer 2018[27] (MP-8; NCT01211405)<br>Ot'alora 2018[28] (MP-12; NCT01793610)<br>Unpublished (MP-4; NCT01958593)<br>Unpublished (MP-9; NCT01689740) |
| Ponte 2021[18]* | Mithoefer 2018[27] (MP-8; NCT01211405)<br>Ot'alora 2018[28] (MP-12; NCT01793610)<br>Unpublished (MP-4; NCT01958593)<br>Unpublished (MP-9; NCT01689740) |

**SI Table 10**: Record of secondary reports that pooled data from multiple clinical trials and were excluded in full-text screening. *Retracted studies or studies with expression of concern due to inclusion of MP-4 data where a previously undisclosed ethical violation occurred.

### Record of Contacting Study Authors

| Study | What info was requested from authors? | What information was provided from authors? |
|---|---|---|
| MP-9 (NCT01689740) | Mean, SD, and N for:<br>● Baseline CAPS-IV and BDI-II<br>● CAPS-IV and BDI-II at end of stage 1 | N/A (no response received) |
| Mitchell 2023 | Mean, SD, and N for:<br>● BDI-II at baseline<br>● BDI-II at T4 | N/A (no response received) |

**SI Table 11**: Record of author contact and responses via email.

### Risk of Bias Assessment Standard Operating Procedure

Standard Operating Procedure for using Cochrane's RoB2 tool. See the *Risk of Bias* section in the supplementary methods for more general details and discussion.

#### Overall Workflow

| **Notes:** |
|---|
| 1. Each reviewer completes the RoB2 tool Excel spreadsheet, using the descriptions in the tool as well as this SOP for guidance.<br>2. Discuss and create a consensus version of the RoB2 tool Excel spreadsheet.<br>3. Create a new spreadsheet from the consensus spreadsheet that includes only overall D1-D5 results for use in R packages to create tables/graphs for RoB.<br>*Always fill out the ITT version of the form, even if the study uses PP analysis.<br><br>*If we override the algorithm in the spreadsheet for any reason, our workflow is to: 1) discuss; 2) document in Notion and the Excel spreadsheet |

#### Domain 1 - Randomization

+ **1.1** - Was the allocation sequence random?
+ **1.2** - Was the allocation sequence concealed until participants were enrolled and assigned to interventions?
+ **1.3** - Did baseline differences between intervention groups suggest a problem with the randomization process?
    + Integrate metapsy tool: If no p-values are provided in the baseline characteristics table for the study you are examining, or if you are uncertain whether the differences are statistically significant, input means, standard deviations, etc. into this tool to determine if baseline differences between intervention groups are significant.
    + Most of the time, the answer to this question will be "no," unless there are substantial differences in the baseline outcome measure between groups. You only need to check the outcome measure (i.e., MADRS, BIDS, CAPS) - there is no reason to think that age or sex imbalance, for example, would severely influence the results.

#### Domain 2 - Deviations

+ *The key assumption we are making for this domain is that it is meant to address major protocol deviations more specifically, not dropout/missingness, which is the concern of Domain 3.*
+ **2.1** - Were participants aware of their assigned intervention during the trial?
    + The answer will probably be "PY" (due to side effects of psychedelics) unless the study actually assesses blinding of participants, and from their assessment you can confidently mark 'Y' if participants were definitely aware of their assigned intervention or 'N' if they were not.
+ **2.2** - Were carers and people delivering the interventions aware of participants' assigned intervention during the trial?
    + The answer will probably be "PY" (due to side effects of psychedelics) unless the study actually assesses blinding of staff and you can confidently mark 'Y' or 'N'.
+ **2.3** - Were there deviations from the intended intervention that arose because of the trial context?
    + Try not to put 'NI' for this even if information is sparse - it actually makes a difference in the algorithm.
    + Things outside of trial context (i.e., participant dropout or restarting medication) usually do not count here. Only count starting medication again if there is a solid reason that one group had a higher rate of starting up medication.
    + An example of what counts as a deviation due to trial context is if one arm received greater levels of psychotherapy compared to the other arm due to safety concerns.
+ **2.4** - Were these deviations likely to have affected the outcome?
    + You can answer 'N' if ITT and PP analyses were consistent.
+ **2.5** - Were these deviations from intended intervention balanced between groups?
+ **2.6** - Was an appropriate analysis used to estimate the effect of assignment to intervention?
    + If it is unspecified whether ITT or PP analysis was used, you can put 'PN'.
+ **2.7** - Was there potential for a substantial impact (on the result) of failure to analyze participants in the group to which they were randomized?
    + Cochrane states for this question that "it is not possible to specify a precise rule." Usually, this question will be PN/N, unless there is evidence of substantial impact.
+ For studies that use PP analysis, but have low/balanced deviations or if the deviations described occur after the primary endpoint that is being assessed, the overall risk of bias for this domain can be overridden to ‘Low’ rather than the algorithm’s assessment of ‘Some concerns’.

#### Domain 3 - Missing Data/Dropout

+ **3.1** - Were data for this outcome available for all, or nearly all, participants randomized?
    + Imputed data still counts as missing data.

        + Calculate: for continuous outcomes, 95% counts as 'nearly all'.
+ **3.2** - Is there evidence that the result was not biased by missing outcome data?
    + If dropouts are low and/or balanced between the two groups, the answer will probably be PY/Y.
    + If dropouts are high and/or unequal between the two groups (especially if related to adverse events/severity of symptoms), and robust sensitivity analyses were not performed to test if there is something different about the groups at baseline or other timepoints, then the answer will probably be PN/N.
+ **3.3** - Could missingness in the outcome depend on its true value?
    + This will be 'PY' if the main reason people dropped out was due to responding poorly (or really well) to treatment.
    + Most of the time, the answer to this question will be 'Y/PY' unless the documented reasons for dropout were all unrelated to health status/outcome.
+ **3.4** - Is it likely that missingness in the outcome depended on its true value?
    + Usually, the answer to this question will be 'PN'. Only put 'Y' for this question unless it is well-documented that missingness in the outcome depended on its true value (i.e., if all the dropouts were due to increased suicidality, or if all participants in the intervention group dropped out because their condition resolved and they no longer felt the need to continue the trial).

#### Domain 4 - Measurements

+ **4.1** - Was the method of measuring the outcome inappropriate?
    + Usually 'N'.
+ **4.2** - Could measurement or ascertainment of the outcome have differed between intervention groups?
    + Usually 'N'.
+ **4.3** - Were outcome assessors aware of the intervention received by study participants?
    + In some cases, it might be necessary to contact study authors, i.e., if the outcome is clinician-administered (i.e., MADRS), and the study does not specify who measured the outcome.
    + Clinician-administered outcomes where blinded third-party or independent study staff raters performed the measurement receive a ‘N’.
    + If the authors do not respond, it is fair to put 'NI' here and 'PY' for 4.4.
    + Self-report outcome measurements get a 'PY' here.
+ **4.4** - Could assessment of the outcome have been influenced by knowledge of intervention received?
    + For self-report, if there is no evidence that participants under-reported, this will be 'PY.'
    + For clinician-administered outcomes with unblinded/functionally unblinded study staff, this will be 'PY'.
+ **4.5** - Is it likely that the assessment of the outcome was influenced by knowledge of intervention received?

    + For self-report, this question will be 'PN' unless there is substantial evidence or reason to believe that participants underreported their symptoms.
    + For clinician-administered outcomes by non-independent study staff, a judgement call must be made based on their level of patient contact, potential for functional unblinding, and overall involvement in the study.
    + For clinician-administered outcomes in an open-label study assessed by explicitly unblinded non-independent study staff, this will be a ‘PY’.

#### Domain 5 - Pre-registration

+ **5.1** - Were the data that produced these results analyzed in accordance with a pre-specified analysis plan that was finalized before unblinded outcome data were available for analysis?
    + Look at the trial protocol/statistical analysis plan if provided.
    + Look at clinicalTrials.gov (making sure to examine the version history).
+ **Is the numerical result being assessed likely to have been selected, on the basis of the results, from...**
+ **5.2** - ...multiple eligible outcome measurements (e.g., scales, definitions, timepoints) within the outcome domain?
+ **5.3** - ...multiple eligible analyses of the data?
